# Household transmission of SARS-CoV-2 from adult index cases living with and without HIV in South Africa, 2020-2021: a case-ascertained, prospective observational household transmission study

**DOI:** 10.1101/2022.04.08.22273160

**Authors:** Jackie Kleynhans, Sibongile Walaza, Neil A. Martinson, Mzimasi Neti, Anne von Gottberg, Jinal N. Bhiman, Dylan Toi, Daniel G. Amoako, Amelia Buys, Kedibone Ndlangisa, Nicole Wolter, Leisha Genade, Lucia Maloma, Juanita Chewparsad, Limakatso Lebina, Linda de Gouveia, Retshidisitswe Kotane, Stefano Tempia, Cheryl Cohen

## Abstract

**Background:** In South Africa 19% of the adult population aged 15-49 years are living with HIV (LWH). Few data on the influence of HIV on SARS-CoV-2 household transmission are available.

**Methods:** We performed a case-ascertained, prospective household transmission study of symptomatic index SARS-CoV-2 cases LWH and HIV-uninfected adults and their contacts in South Africa. Households were followed up thrice weekly for 6 weeks to collect nasal swabs for SARS-CoV-2 testing. We estimated household cumulative infection risk (HCIR), duration of SARS-CoV-2 positivity (at cycle threshold value<30 as proxy for high viral load), and assessed associated factors.

**Results:** We recruited 131 index cases and 457 household contacts. HCIR was 59% (220/373); not differing by index HIV status (60% [50/83] in cases LWH vs 58% [173/293] in HIV-uninfected cases, OR 1.0, 95%CI 0.4-2.3). HCIR increased with index case age (35-59 years: aOR 3.4 95%CI 1.5-7.8 and ≥60 years: aOR 3.1, 95%CI 1.0-10.1) compared to 18-34 years, and contacts’ age, 13-17 years (aOR 7.1, 95%CI 1.5-33.9) and 18-34 years (aOR 4.4, 95%CI 1.0-18.4) compared to <5 years. Mean positivity duration at high viral load was 7 days (range 2-28), with longer positivity in cases LWH (aHR 0.3, 95%CI 0.1-0.7).

**Conclusions:** HIV-infection was not associated with higher HCIR, but cases LWH had longer positivity duration at high viral load. Adults aged >35 years were more likely to transmit, and individuals aged 13-34 to acquire SARS-CoV-2 in the household. Health services must maintain HIV testing with initiation of antiretroviral therapy for those HIV-infected.

**Summary:** In this case-ascertained, prospective household transmission study, household cumulative infection risk was 59% from symptomatic SARS-CoV-2 index cases, not differing based on index HIV status. Index cases living with HIV were positive for SARS-CoV-2 for longer at higher viral loads.

## Introduction

At end of January 2022, South Africa had reported 3.6 million coronavirus disease 2019 (COVID-19) cases and 94.3 thousand deaths; the highest reported from Africa, likely due to the highest testing capacity.[1, 2] South Africa experienced four severe acute respiratory syndrome coronavirus 2 (SARS-CoV-2) waves of infection: the first dominated by ancestral variants, followed by Beta (B.1.351), Delta (B.1.617.2) and Omicron (B.1.1.529) variant dominated waves, respectively.[3]

Although incident HIV infections and AIDS-related deaths from 2010 to 2019 in South Africa reduced by 53% and 61%, respectively, the burden of HIV in South Africa is still high, with an estimated 19% of the adult population aged 15-49 living with HIV (LWH); the fourth highest in Sub-Saharan Africa.[4] In 2019, South Africa had an estimated 186,000 new HIV infections and 68,000 adult deaths were HIV/AIDS-related.[4] However, South Africa has managed to attain two of the three 90-90-90 goals: an estimated 92% of South Africans of all ages know their HIV status, 75% of whom are on antiretroviral treatment (ART), and 92% of individuals on treatment are virally suppressed.[3]

Few studies report the influence of HIV infection on SARS-CoV-2 transmission, duration of shedding and clinical severity, with most data available from high income countries and little evidence from sub-Saharan Africa where most people LWH reside.[4] Evidence from multiple studies suggests that people LWH are at greater risk for hospitalisation[5-8] and death[7-11] when infected with SARS-CoV-2, but that SARS-CoV-2 prevalence is similar between people LWH and HIV-uninfected individuals,[12] Risk for hospitalisation and death increases with a decline in CD4+ T cells.[6, 7, 11] Limited data are available on the role of HIV in the transmission of SARS-CoV-2. One study from a South African household cohort showed no increase in household transmission from or acquisition of SARS-CoV-2 infection in people LWH.[2] People LWH with severe COVID-19 who are not virally suppressed shed SARS-CoV-2 for longer periods,[2, 13] which could also lead to increased secondary transmission.

Case-ascertained household transmission studies can assist in better understanding risk factors to transmit and acquire SARS-CoV-2, the proportion of household contacts that become infected and that develop symptoms, the duration of shedding and the serial interval, but few studies have been published thus far, especially from Africa. A recent meta-analysis including 126 studies, estimated a 36% household secondary attack for studies done in 2021, and 16% for studies done in 2020.[14]

We aimed to assess household cumulative infection risk, duration of SARS-CoV-2 positivity (episode duration), serial interval and factors associated with these measures in households with SARS-CoV-2 index cases LWH and HIV-uninfected.

## Methods

We conducted a case-ascertained, prospective observational household transmission study of household contacts of symptomatic adult index SARS-CoV-2 cases LWH and HIV-uninfected at two sites in South Africa, Klerksdorp (North West Province) and Soweto (Gauteng Province). Sample size calculation is detailed in the supplementary methods. The total sample size was 264 and 176 contacts from households with an HIV-uninfected and HIV-infected index case, respectively. This was based on assuming 40% HIV prevalence among index cases, HCIR of 10% and 20% in household members exposed to an HIV-uninfected and HIV-infected index case, respectively, for a 95% confidence interval and 80% power.

### Screening for index cases

Screening procedures are detailed in the supplementary methods. In short, nasopharyngeal swabs were collected from clinic attendees aged ≥18 years with symptom onset ≤5 days prior to screening and tested for SARS-CoV-2 RNA using real-time reverse transcription polymerase chain reaction (rRT-PCR).

### Household enrolment

We approached households of individuals who tested positive for SARS-CoV-2, with symptom onset less than 7 days prior, with no household members reporting symptoms in the 14 days prior to index screening. Households with ≥3 eligible members, of whom ≥70% consented to participate were enrolled. We collected information on household characteristics, demographics, underlying medical conditions, and vaccination information. Households that withdrew within 10 days from index symptom start date were excluded from the analysis.

### Index and household follow-up

We visited households three times a week for 6 weeks to collect a nasal swab, presence of symptoms and healthcare seeking behaviour from all consenting household members. At the first and last study visits, clotted blood was collected for serological testing. All data collected during screening and follow up visits were captured in real-time using tablets onto Research Electronic Data Capture (REDCap) databases hosted at the University of the Witwatersrand[15]. Follow-up continued to 11 August 2021 and 28 September 2021 in Klerksdorp and Soweto respectively.

### SARS-CoV-2 detection

Nasopharyngeal (screening) and nasal (follow-up) specimens were tested for the E, RdRp and N SARS-CoV-2 genes by rRT-PCR, using the Allplex(tm) 2019-nCoV kit (Seegene Inc., Seoul, South Korea) according to manufacturer instructions. Specimens were considered positive for SARS-CoV-2 if the cycle threshold (C_t_) value was below 40 for any of the gene targets.

### SARS-CoV-2 variants

We characterized the first SARS-CoV-2 positive specimen for each participant on the Allplex(tm) SARS-CoV-2 Variants I and II PCR assays (Seegene Inc., Seoul, Korea) and through full genome sequencing on the Ion Torrent Genexus platform (Thermo Fisher Scientific, USA). We classified the infection episodes as an Alpha, Beta, Delta, non-Alpha/Beta/Delta or unknown variant as detailed in supplementary methods.

### Serology

We used an in-house ELISA to detect antibodies against SARS-CoV-2 spike protein[16] and nucleocapsid protein using the Roche Elecsys anti-SARS-CoV-2 assay. Individuals were considered seropositive if they tested positive on either one of the assays performed.

### Statistical analysis

Definitions used are defined in Table 1. To assess factors associated with HCIR and minimum Ct value ≤25 during secondary cases episode we used logistic regression accounting for within site and household clustering using a mixed effects hierarchical regression model. To assess factors associated with a time to event analysis (serial interval and episode duration in index cases), we used a multilevel mixed-effects survival model with Weibull accelerated failure time analysis, controlling for site and household clustering in serial interval analysis, and site clustering in the episode duration analysis. Episode duration was assessed at any Ct value (<40), and as rRT-PCR positivity with a Ct value for any target with a Ct<30 (proxy for high viral load). We first assessed co-variates on univariate analysis, including all with p<0.2 in the multivariable analysis. We performed backwards elimination and kept all variables with p<0.05 in the final model, except for those included a priori. We included site, SARS-CoV-2 variant and index immune suppression related to HIV status (defined as CD4+ T cell count <200 cells/ml) in the HCIR and episode duration models; and site in the serial interval analysis. Variant type and immune suppression status was not included a priori in the serial interval analysis due to small numbers, and no predictors were included a prior in the minimum Ct value analysis.

**Table 1.**
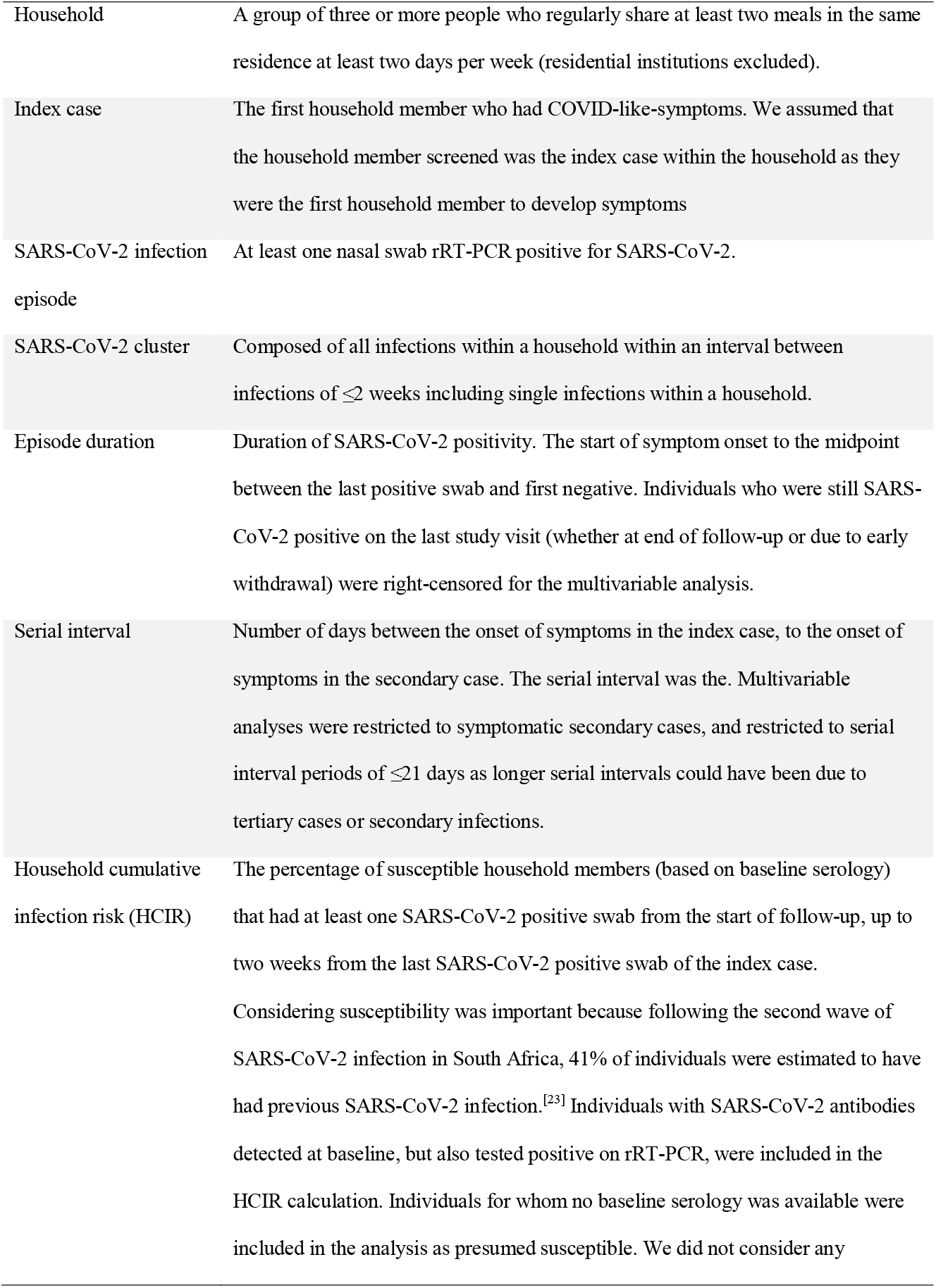
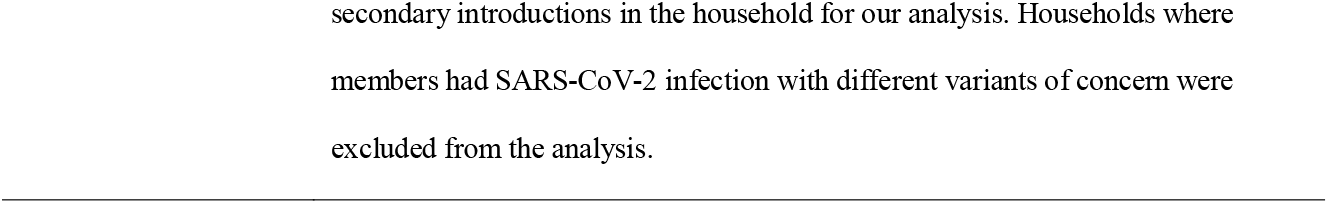
Definitions

|  |  |
| --- | --- |
| Household | A group of three or more people who regularly share at least two meals in the same residence at least two days per week (residential institutions excluded). |
| Index case | The first household member who had COVID-like-symptoms. We assumed that the household member screened was the index case within the household as they were the first household member to develop symptoms |
| SARS-CoV-2 infection episode | At least one nasal swab rRT-PCR positive for SARS-CoV-2. |
| SARS-CoV-2 cluster | Composed of all infections within a household within an interval between infections of $\leq 2$ weeks including single infections within a household. |
| Episode duration | Duration of SARS-CoV-2 positivity. The start of symptom onset to the midpoint between the last positive swab and first negative. Individuals who were still SARS-CoV-2 positive on the last study visit (whether at end of follow-up or due to early withdrawal) were right-censored for the multivariable analysis. |
| Serial interval | Number of days between the onset of symptoms in the index case, to the onset of symptoms in the secondary case. The serial interval was the. Multivariable analyses were restricted to symptomatic secondary cases, and restricted to serial interval periods of $\leq 21$ days as longer serial intervals could have been due to tertiary cases or secondary infections. |
| Household cumulative infection risk (HCIR) | The percentage of susceptible household members (based on baseline serology) that had at least one SARS-CoV-2 positive swab from the start of follow-up, up to two weeks from the last SARS-CoV-2 positive swab of the index case.<br>Considering susceptibility was important because following the second wave of SARS-CoV-2 infection in South Africa, 41% of individuals were estimated to have had previous SARS-CoV-2 infection.<sup>[23]</sup> Individuals with SARS-CoV-2 antibodies detected at baseline, but also tested positive on rRT-PCR, were included in the HCIR calculation. Individuals for whom no baseline serology was available were included in the analysis as presumed susceptible. We did not consider any |

### Sensitivity analysis

To assess the influence of loss to follow-up on the results, we performed a sensitivity analysis including only households where 65% of household members completed 65% of follow-up visits in the first three weeks of follow-up. As a second sensitivity analysis, to explore the effect of likely previous SARS-CoV-2 infection, we also considered all household members irrespective of baseline serology as susceptible contacts in the HCIR analysis.

### Ethics

The study protocol was approved by the University of Witwatersrand Human Research Ethics Committee (Reference M2008114). Participants in household follow-up received grocery store vouchers of USD 3 per visit to compensate for time required for specimen collection and interview.

## Results

### Screening, enrolment and follow-up

From 2 October 2020 to 30 September 2021, we screened 1,531 clinic attendees for SARS-CoV-2, of whom 18% (277) tested positive on rRT-PCR. Of those testing positive for SARS-CoV-2 who met eligibility criteria for household enrolment (n=277), 143 (52%) households were approached and 131 (92%) were enrolled and included in the analysis. The final cohort consisted of 131 index cases and 457 household contacts (Figure 1).

**Figure 1.**
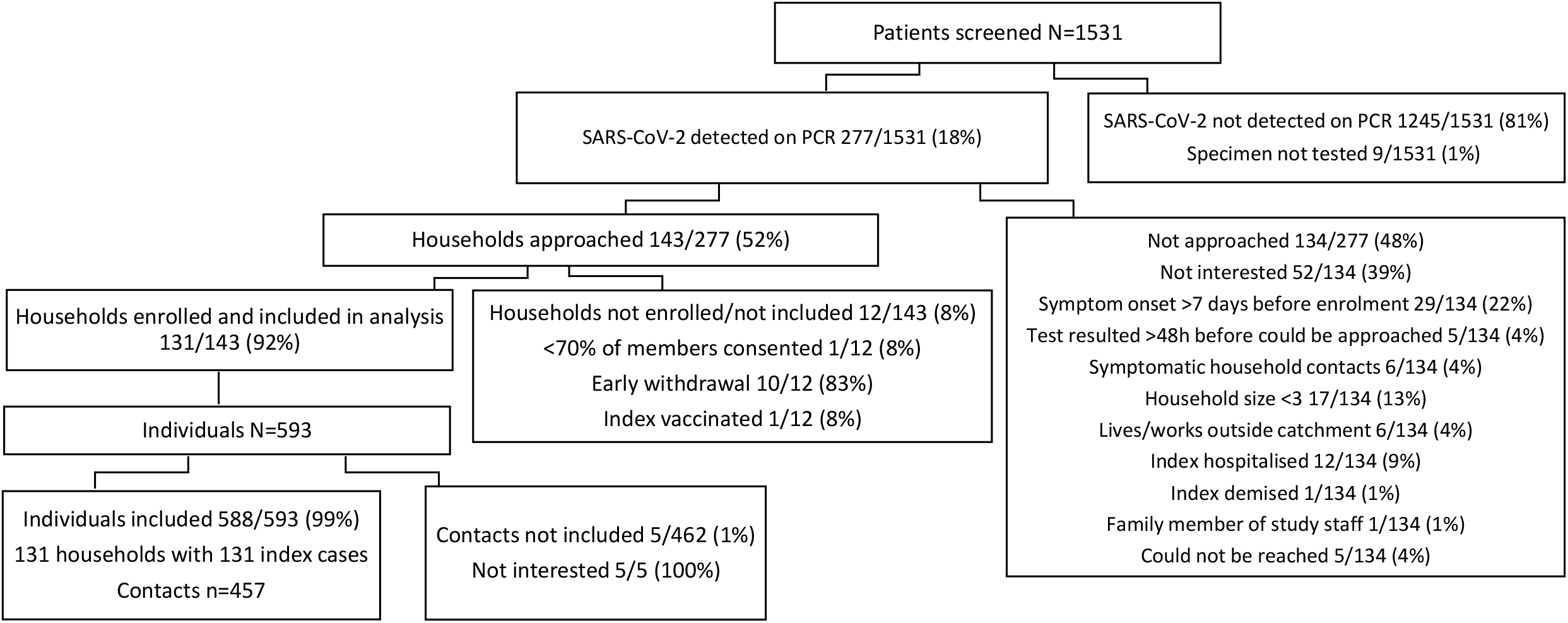
Adult SARS-CoV-2 index cases and household contacts enrolled, Klerksdorp and Soweto, South Africa, 2020-2021.

**Figure 2.**
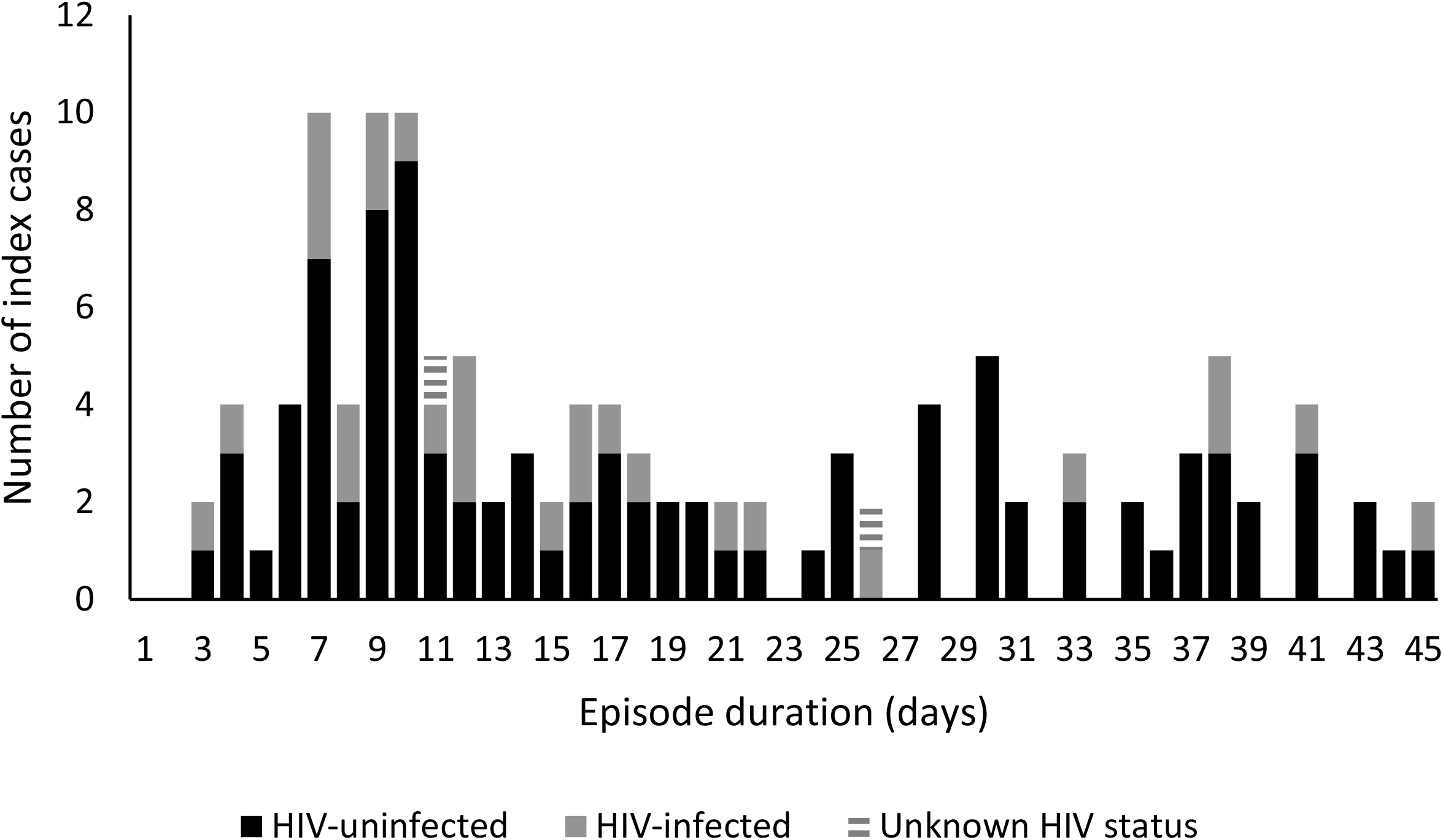
SARS-CoV-2 episode duration in HIV-infected and HIV-uninfected index cases, Klerksdorp and Soweto, South Africa, 2020-2021, (n=123). Excludes those positive at last specimen collected.

Twenty-one percent (28/131) of index cases were LWH, and two index cases initially agreed but then refused HIV testing after enrolment. The majority (93/131, 71%) of index cases and contacts (265/457, 58%) were female. The mean household size was 5 individuals (range 3 – 10) and 67% of households reported >2 people sleeping in the same room (Table 2).

**Table 2.**
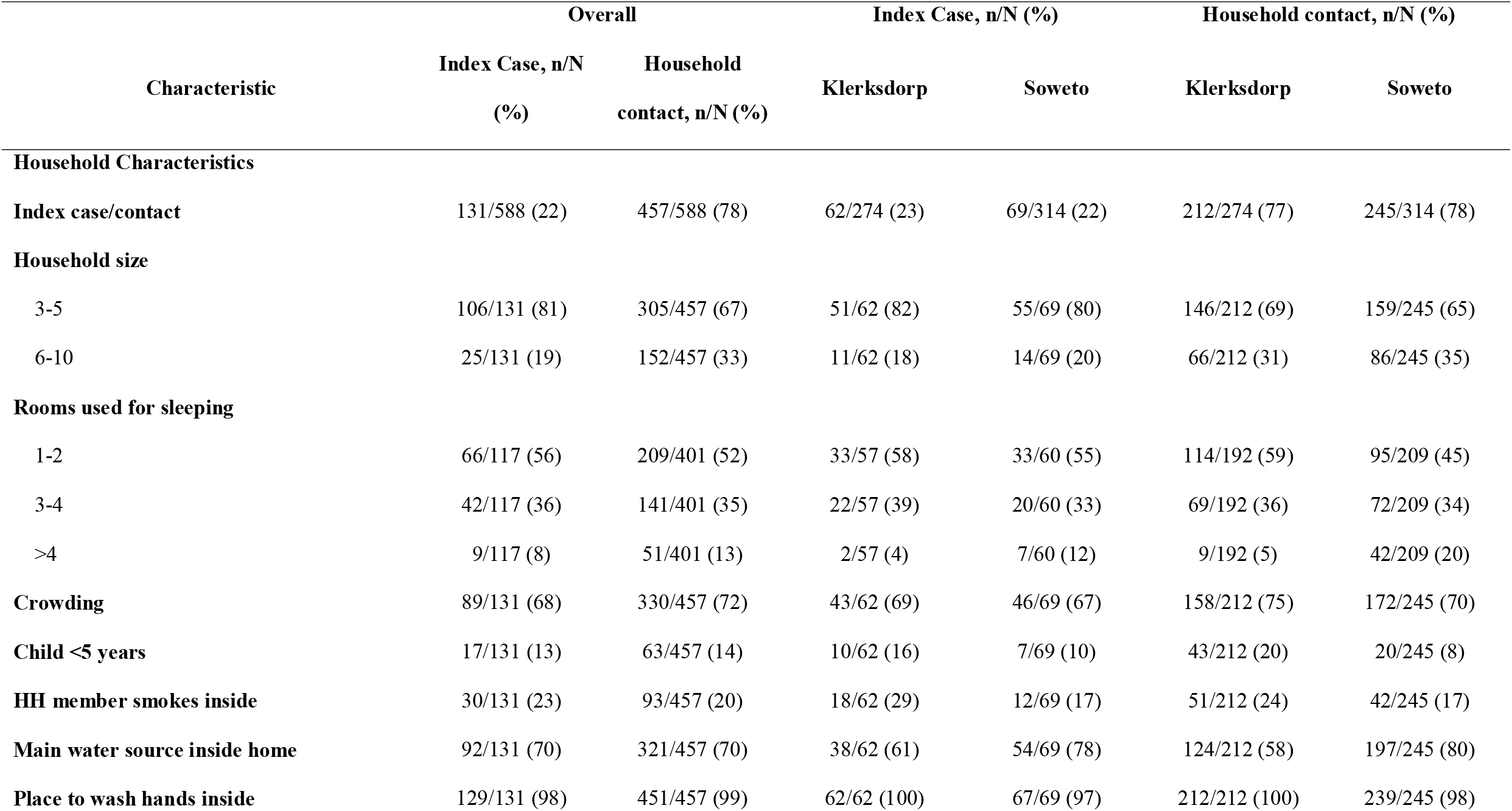
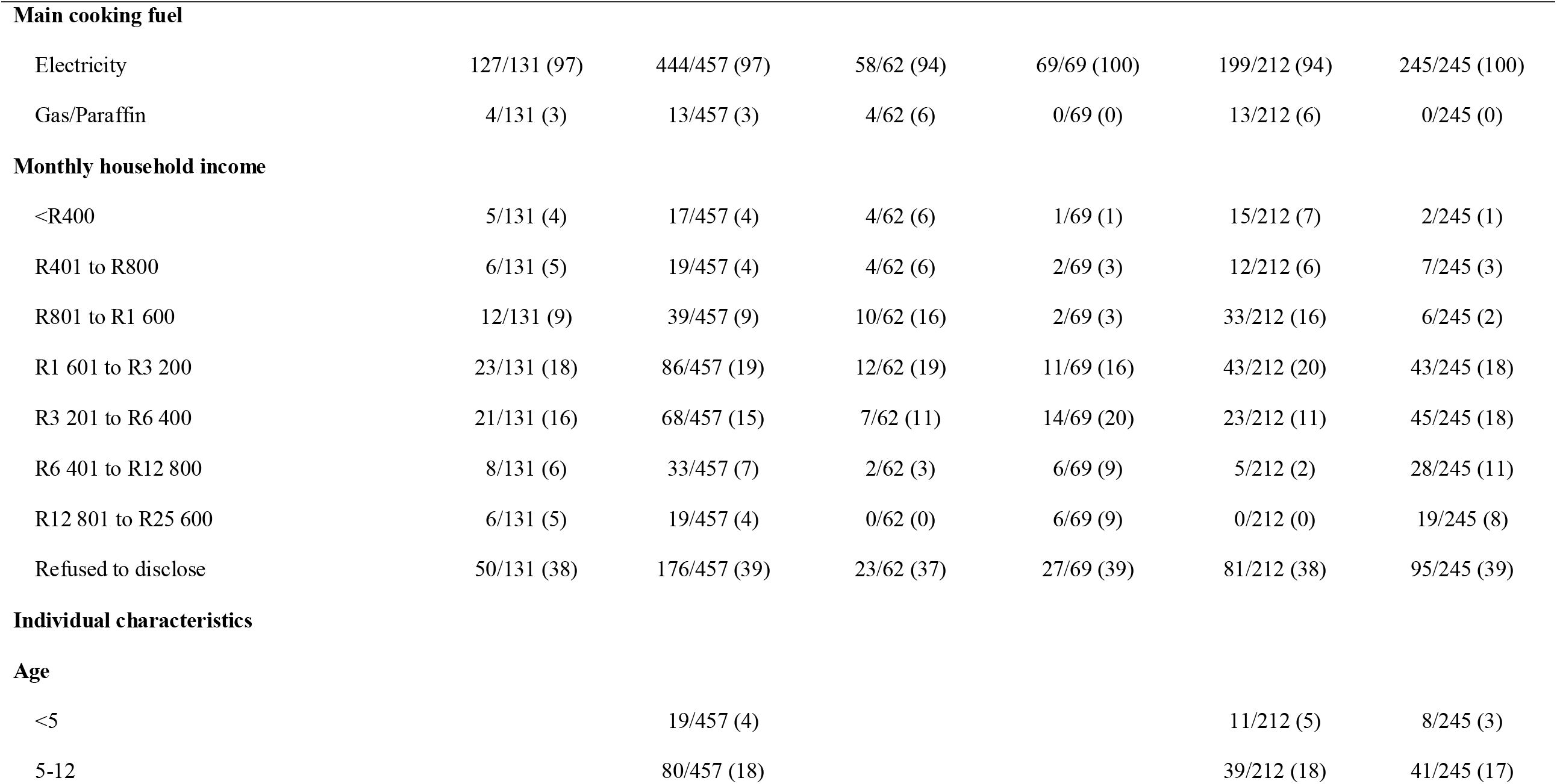
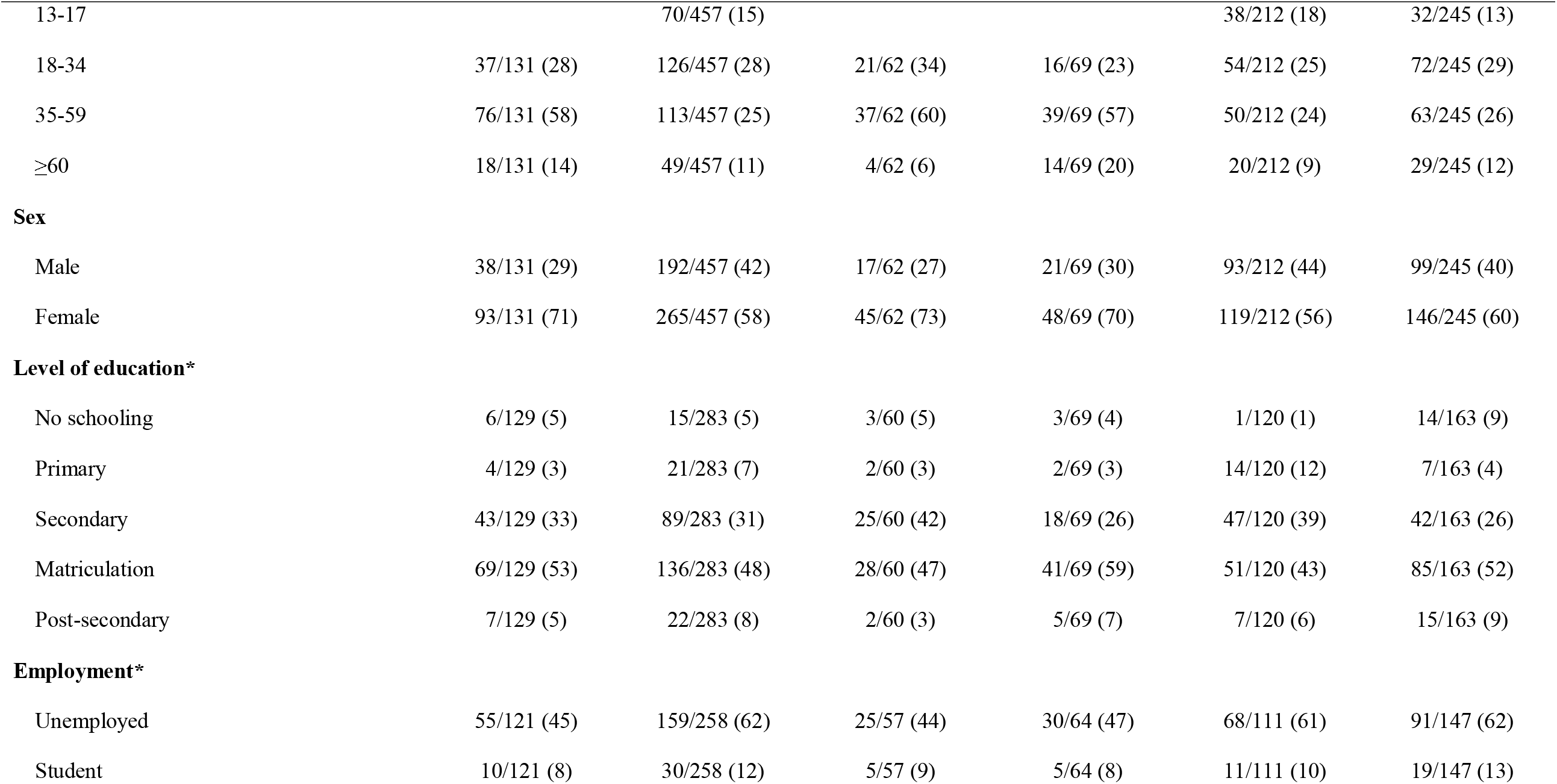
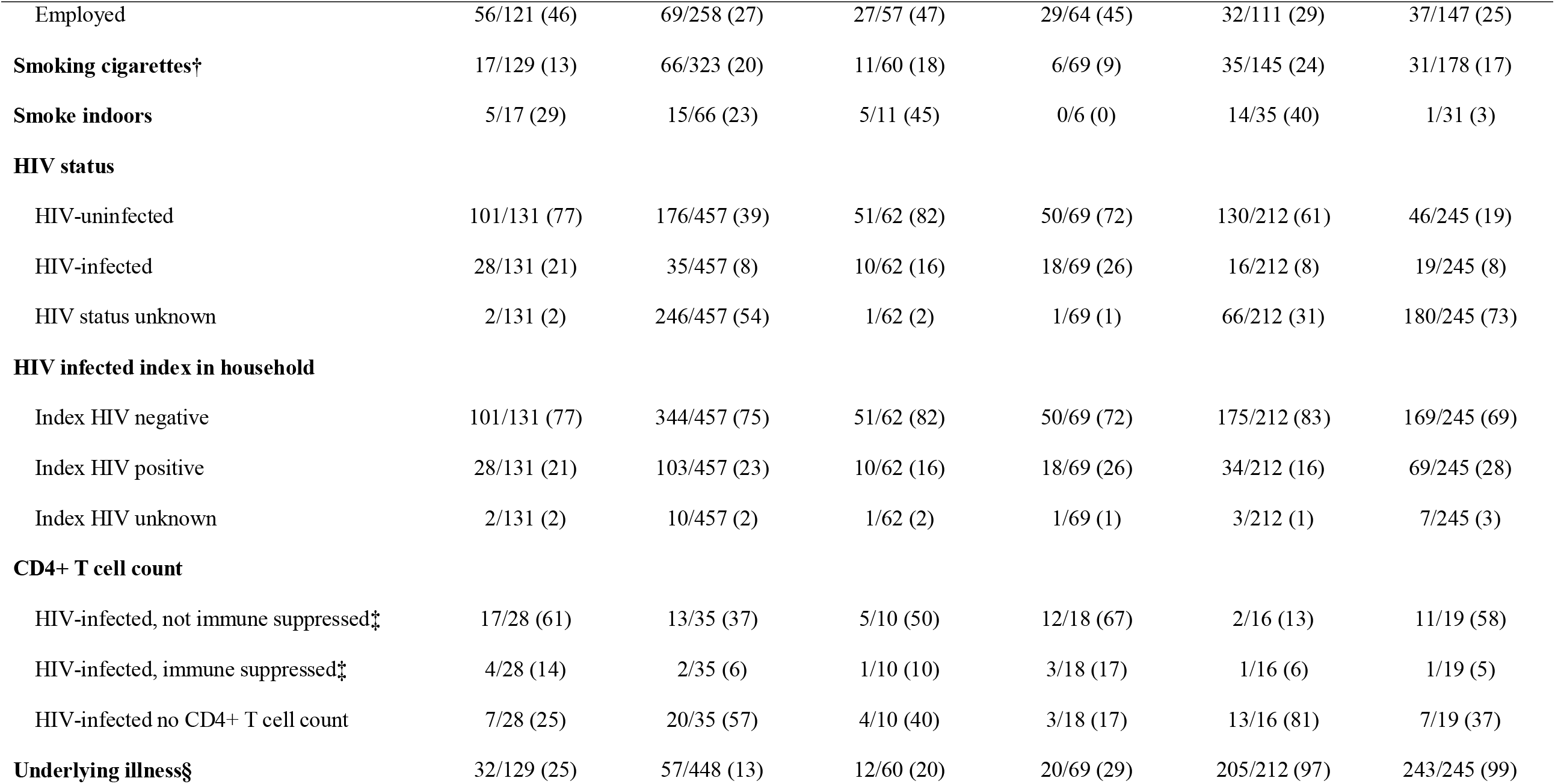
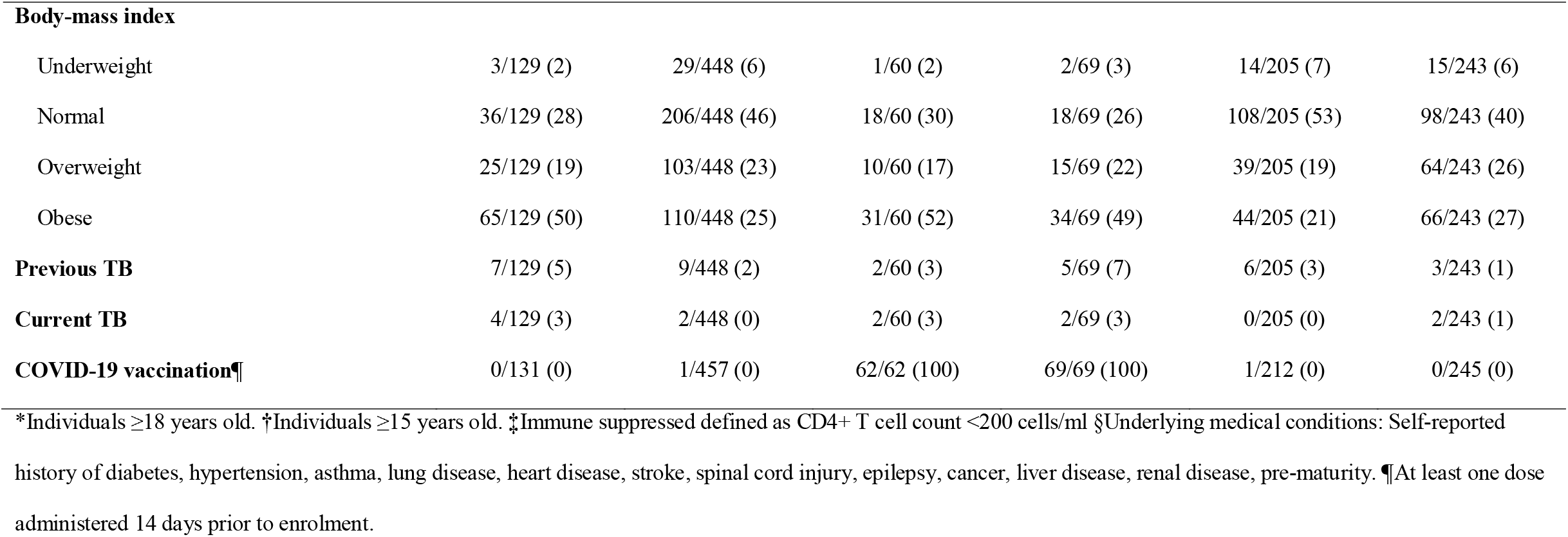
Baseline characteristics of SARS-CoV-2 index cases (n=131) and their household contacts (n=457), Klerksdorp and Soweto, South Africa, September 2020 – October 2021.

Of a potential 10,584 study visits, we completed 8,509 (80%) visits, and detected SARS-CoV-2 in 17% (1,454/8,352) of nasal swabs collected (Supplementary Figure 1).

### Secondary SARS-CoV-2 cases

We diagnosed 232 laboratory-confirmed secondary cases from the 457 contacts linked to the 131 index cases. One third (69/232) of secondary cases reported at least one symptom during their SARS-CoV-2 episode, with contacts reporting between one to nine symptoms, and an average of three symptoms during the episode. The mean duration of symptoms in secondary cases was 11 days (range 4-40 days). The three most common symptoms reported in contacts were cough (45/69, 65%), headache (31/67, 46%) and self-reported or measured fever ≥38°C (27/69, 39%).

### Household cumulative infection risk

Of 131 households, we excluded eight (6%) households from the HCIR analysis: four (17 contacts) had SARS-CoV-2 clusters with more than one variant detected, and in three (8 contacts) all contacts were seropositive at baseline with no PCR-confirmed SARS-CoV-2 infection during follow-up. An additional 63 contacts were excluded from this analysis because they had prevalent SARS-CoV-2 antibodies at baseline, and no SARS-CoV-2 detection during follow-up. We therefore included 124/131 (95%) index cases with 373/436 (86%) contacts for this analysis.

The HCIR was 59% (220/373) overall. The mean number of household contacts testing positive for SARS-CoV-2 following the index episode was 2 and ranged from 0 to 7. In the univariate analysis, the HCIR was similar in households with HIV-uninfected index cases (58%, 173/293), compared to households where the index was LWH (60%, 50/83, OR 1.0, 95%CI 0.4-2.3).

On multivariable analysis, after adjusting for site and immune suppression, factors significantly associated with household transmission were index case aged 35-59 years (aOR 3.4 95% CI 1.5-7.8) and ≥60 years (aOR 3.1, 95%CI 1.0-10.1) compared to 18-34 years; index cases with a Ct value <25 at any point in the episode (aOR 5.6, 95%CI 1.6-17.6) and 25-35 (aOR 7.5, 95%CI 2.2-26.0) compared to Ct >35; infection with the Delta variant compared to Beta (aOR 4.6, 95%CI 1.5-14.4); index cases that were underweight (aOR 0.1, 95%CI 0.0-0.8) and obese (aOR 0.4 95% CI 0.2-0.9), compared to normal weight (Figure 4, Supplementary table 1). Among index cases LWH, individuals with immune suppression (CD4+ T cell count <200 cells/ml), had a higher HCIR (62%, 8/13) compared to index cases not immune suppressed (58%, 30/52), but this was not statistically significant on multivariable analysis (aOR 2.5, 95%CI 0.4-15.3). Contact age 13-17 years (aOR 7.1, 95%CI 1.5-33.9) and 18-34 years (aOR 4.4, 95%CI 1.0-18.4) compared to <5 years and contacts not currently smoking (aOR 3.9, 95%CI 1.7-9.0) were associated with higher HCIR.

**Figure 3.**
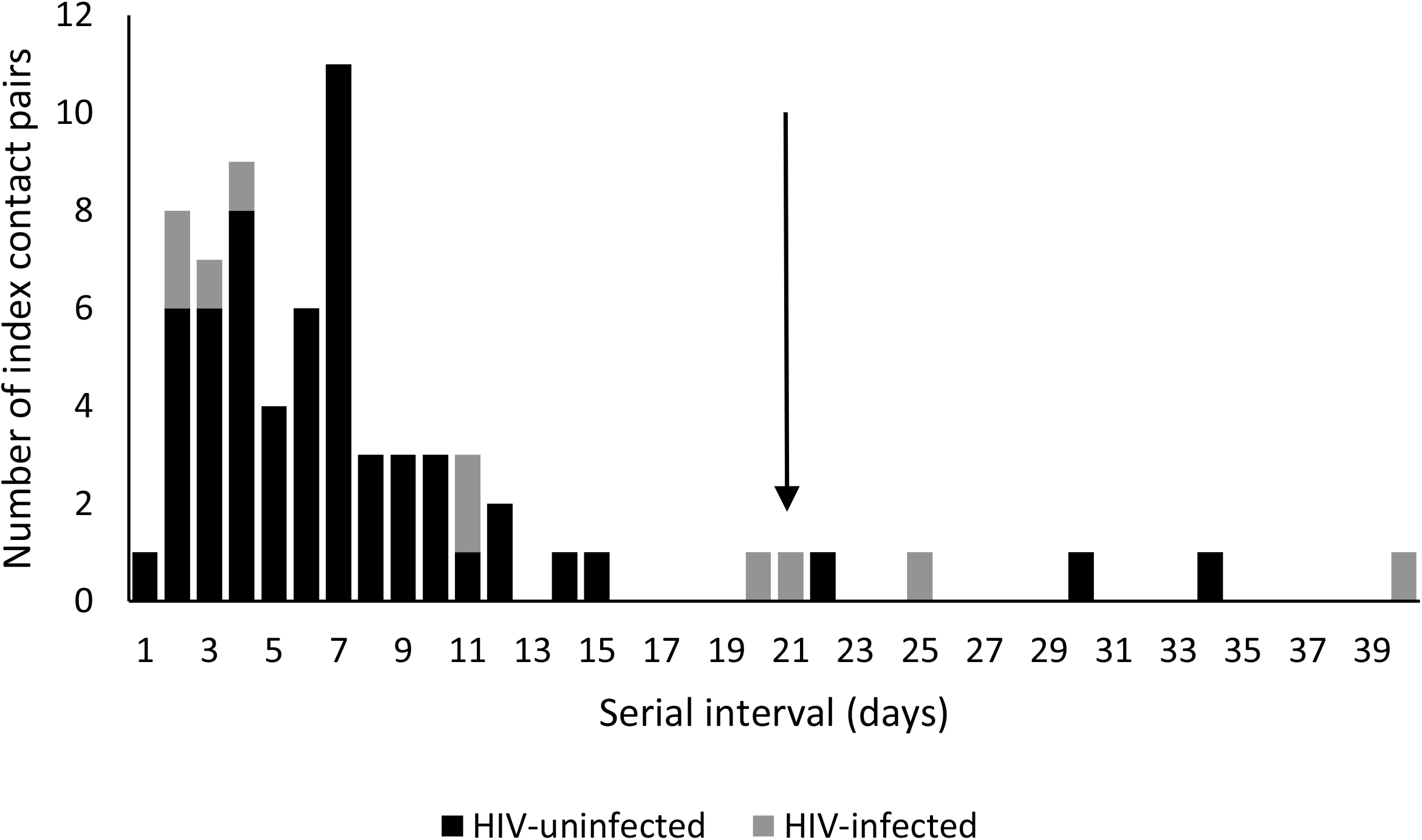
Interval between onset of symptoms in the index case and onset of symptoms in household contacts with SARS-CoV-2 (serial interval) by HIV-infection status of the index case, Klerksdorp and Soweto, South Africa, 2020-2021, (n=69). Arrow indicates cut-off for inclusion in analysis for factors associated with serial interval duration

**Figure 4.**
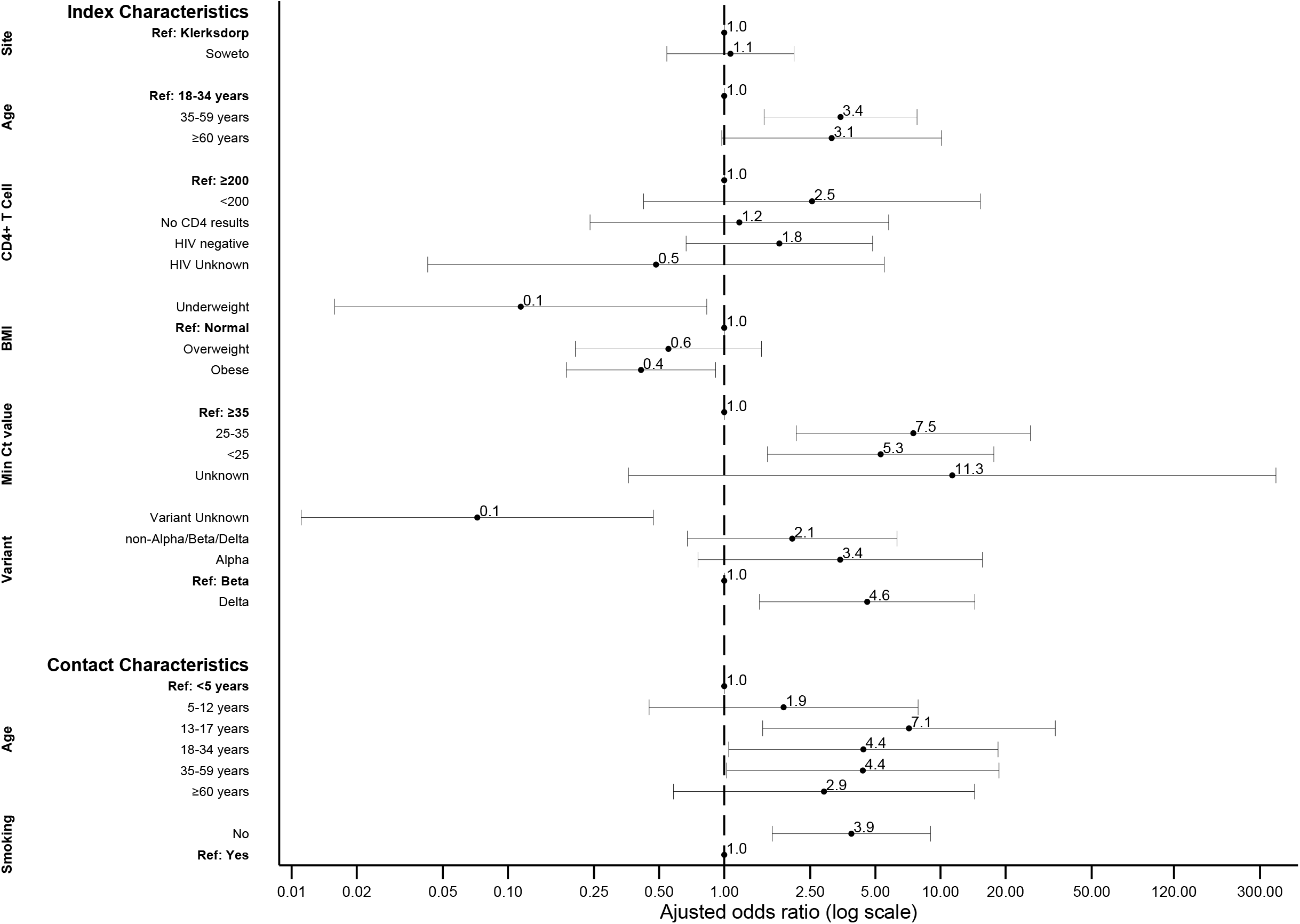
Factors associated with SARS-CoV-2 household transmission in index cases and acquisition in household contacts on multivariable logistic regression, Klerksdorp and Soweto, South Africa, 2020-2021, (n=373).

### Episode duration

We right-censored 6% of the (8/131) index cases who were SARS-CoV-2 positive on their last specimen collected at the end of follow-up (n=5) or at withdrawal (n=3). When including all 131 index cases, the mean episode duration for index cases was 20 days and ranged from 3 to 47 days. When excluding the eight right-censored individuals (n=123), the mean episode duration for index cases was 19 days and ranged from 3 to 45 days. The mean episode duration was similar for HIV-uninfected index cases (20 days, range 3-45) compared to index cases LWH (17 days, range 3-45, HR 0.8, 95%CI 0.5-1.2).

On multivariable analysis, factors associated with a longer episode duration was Soweto site (aHR 0.5, 95%CI 0.3-0.7), being 35-59 years old (aHR 0.4, 95%CI 0.2-0.6) and ≥60 years (aHR 0.2, 95%CI 0.1-0.5) compared to 18-34 years, Ct values <25 (aHR 0.3, 95%CI 0.2-0.6) compared to >35 (Table 3) and being seropositive at the end of follow-up (aHR 0.1, 95%CI 0.0-0.3) compared to seronegative. Individuals infected with the Delta variant (aHR 2.4, 95%CI 1.1-5.0) and a non-variant of concern (aHR 2.5, 95%CI 1.3-4.7), compared to Beta, and those who were underweight (aHR 5.3, 95%CI 1.4-20.0) or obese (aHR 1.9, 95%CI 1.1-3.1) compared to a normal BMI, had shorter episode durations (Figure 5, Supplementary table 2).

**Figure 5.**
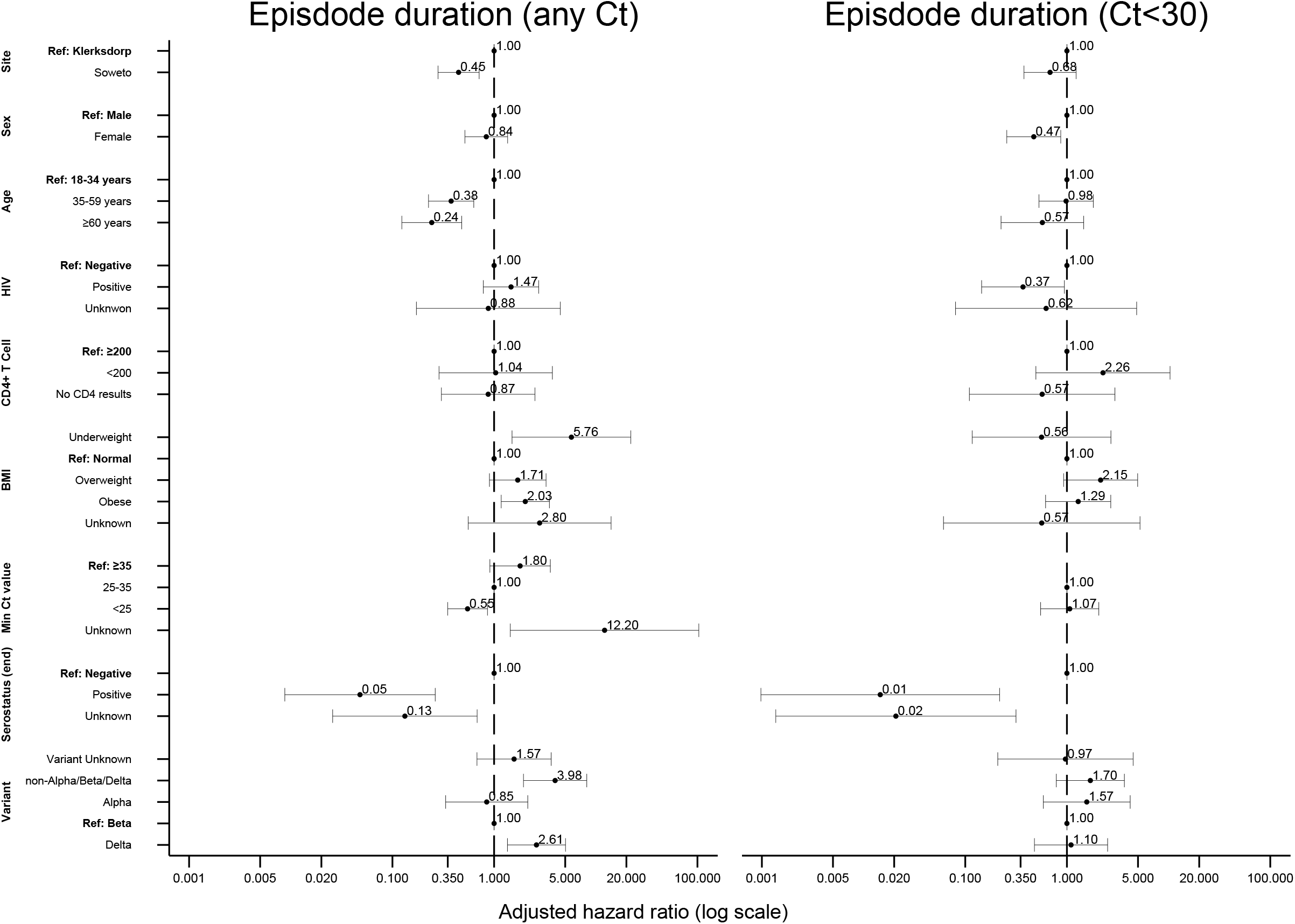
Factors associated with SARS-CoV-2 episode duration (irrespective of Ct value left, Ct<30 right) in index cases on Weibull accelerated failure time regression, Klerksdorp and Soweto, South Africa, 2020-2021, (n=123).

Eighty-eight (67%) of index cases had at least one specimen with at least one target with a Ct<30. The mean episode duration with Ct<30 was 7 days (range 2-17). On multivariable analysis, factors associated with a longer episode duration considering only specimens with at least one target with a Ct<30 was female sex (aHR 0.5, 95%CI 0.3-0.9), LWH (aHR 0.4, 95%CI 0.1-0.9) and being seropositive at the end of follow-up (aHR 0.01, 95%CI 0.001-0.2) compared to seronegative (Figure 5, Supplementary table 3).

### Serial interval

We excluded 5 index contact pairs from the analysis where the serial interval was >21 days; three with an HIV-uninfected contact and two pairs where the index was LWH. The mean serial interval for index cases and symptomatic contact pairs was 6 days and ranged from 1 to 20 days. The mean serial interval for HIV-uninfected index cases was 6 days (range 1-15) and for index cases LWH was 8 days (range 2-20).

On multivariable analysis, pairs with contacts aged 35-59 years (aHR 0.3, 95%CI 0.1-0.9) and aged ≥60 years (aHR 0.2, 95%CI 0.0-0.8), compared to being aged 18-34 years, and where the contact was LWH (aHR 0.1, 95%CI 0.0-0.8) compared to being HIV-uninfected had longer serial intervals (Figure 6, Supplementary table 4).

**Figure 6.**
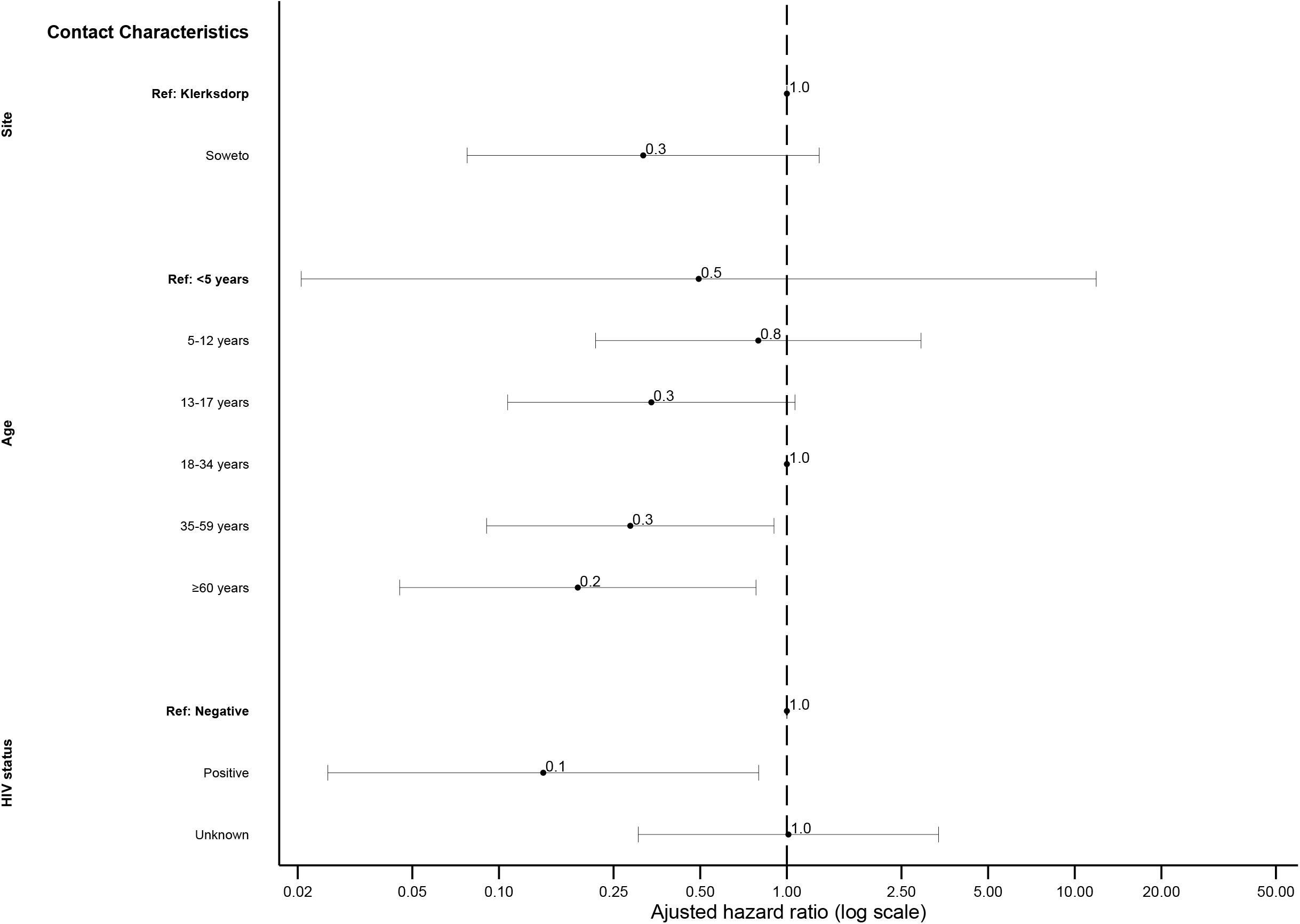
Factors associated with serial interval of SARS-CoV-2 in cases with symptomatic illness on Weibull accelerated failure time regression, Klerksdorp and Soweto, South Africa, 2020-2021, (n=62).

### Factors associated with low minimum Ct value in secondary cases

Of the 232 laboratory-confirmed secondary cases diagnosed, 46% had at least one SARS-CoV-2 positive sample with a Ct value for any target ≤25 (proxy for higher viral load). On multivariable analysis, contacts with underlying illness (aOR 3.7, 95%CI 1.1-12.9), that reported any symptoms (aOR 3.2, 95%CI 1.2-8.8), that were seronegative at the start of follow-up (aOR 9.4, 95%CI 3.7-23.5), seropositive at the end of follow-up (aOR 18.2, 95%CI 3.4-95.9), and infected with the Delta variant (aOR 4.6, 95%CI 1.2-17.9) compared to Beta, were more likely to have had at least one SARS-CoV-2 positive sample with a Ct value for any target ≤25 (Supplementary table 5).

### Out-patient consultations, hospitalisations and deaths

The COVID-19-related hospitalisation rate was 6% (6/131) in index cases and 1% (5/457) in contacts (Supplementary Table 6), with one death each of an index case and contact. In addition to the contacts admitted, only 2 other contacts reported outpatient consultation visits related to the SARS-CoV-2 episode.

### Sensitivity analysis

In a sensitivity analysis in which we did not exclude individuals that were seropositive at baseline with no PCR confirmed SARS-CoV-2 infection during follow-up, the HCIR was 51% (220/436) overall, 46% (95/205) and 54% (125/231) in Klerksdorp and Soweto respectively. We still found similar factors associated with HCIR (Supplementary Table 2), with HCIR in households with an index LWH being the same as in households where the index was HIV-uninfected (49%). Furthermore, when only including households where 65% of members completed 65% of visits of their first three weeks of follow-up, we included 112 index cases with 342 contacts. Factors associated with HCIR, episode duration and serial interval were similar to what we observed in the main analysis (Supplementary Tables 7-9)

## Discussion

We performed a case-ascertained, prospective observational household transmission study for SARS-CoV-2 in South Africa, including 131 index cases, 28 of whom were LWH; and 457 household contacts. The HCIR was similar in index cases LWH and HIV-uninfected index cases. Episode duration at high viral load was longer in index cases LWH and serial interval was longer in contacts LWH. We observed a 59% HCIR, with HCIR being higher in households with older index cases and contacts aged 13-17 years and 18-34 years. HCIR was also higher in households with Delta-infected index cases compared to Beta. Index episode durations were longer in older aged individuals, and in those with a Delta infection compared to Beta. The serial interval was longer if the contact was older and LWH.

HCIR from previous studies have varied greatly based on study design, symptom status of the index timing within the epidemic[17] and SARS-CoV-2 variant.[14] In our study only including symptomatic index cases, we estimated the HCIR at 59%, a higher estimate than the 36% reported from a recent meta-analysis.[14] In a Madagascar study also including both symptomatic and non-symptomatic index cases, HCIR was estimated at 39%.[18] The higher estimate seen in our current study may be influenced by symptom severity, as proposed in previous studies,[19] or the inclusion of only adult index cases, since adult index cases have been shown to result in higher HCIR.[20] In our study, households with index cases older than 35 years were three times more likely to result in higher HCIR compared to when index cases were aged 18-34 years. HCIR was also higher when contacts were aged 13-17 and 18-34, compared to younger than 5 years. Contacts aged 13-18 years were also associated with higher HCIR in the South African PHIRST-C study,[2] although studies from earlier in the pandemic showed higher attack rates in elderly household members.[19, 21] This may be related to the shift in age-distribution of cases from the older population to younger individuals with progression of the pandemic.[2, 22, 23] As seen previously,[2, 24, 25] we also observed higher secondary attack rates where the minimum rRT-PCR cycle threshold was lower for the index case, which could be considered a proxy for higher viral load.

We observed no difference in HCIR in households with index cases living with and without HIV. However, we observed a signal for higher HCIR in PLWH who were immune suppressed, but this association was not statistically significant possibly due to low numbers (n=14) of included immunosuppressed index cases LWH. This would fit with previous studies finding that immunocompromised PLWH that shed virus for longer[2, 13] allowing longer opportunity for secondary infections.

The mean episode duration of 19 days, was higher than the 11 days reported from the household cohort study from South Africa[2] but more similar to the 18 days estimate from a meta-analysis for viral shedding time.[26] This may be due to our analysis being limited to symptomatic individuals, which was shown to be associated with longer episode duration.[2, 26] Furthermore, our episode duration estimates could also have been biased to shorter durations due to the right-censoring of individuals still positive for SARS-CoV-2 by the end of follow up. However, episode duration in this study is also higher than studies on hospitalised South African patients where the median episode duration was 13 days[13] and 11 days. Previous studies from South Africa in the community and in hospitals found that immunocompromised people LWH shed SARS-CoV-2 for longer.[2, 13] While we did not find overall longer shedding in people LWH, when considering detection at a Ct<30 (proxy for high viral load), we also observed people LWH had longer episode durations.

Obesity has been shown to be associated with increased episode duration,[13] of which we observed the inverse with individuals of normal weight having longer episode durations, the reason for this is unclear. After adjusting for confounders on multivariable analysis, individuals infected with the Delta variant had shorter episode durations than those infected with Beta, while episode duration did not differ based on variant when considering detection at a Ct<30. In contrast, households with Delta infections had higher HCIR. Longer episode duration does not necessary indicate shedding of live virus; and higher HCIR with Delta might be related to higher viral loads[27] and increased infectiousness.[28] We did observe lower minimum Ct values (proxy for higher viral load) in secondary cases infected with Delta compared to Beta. The shorter episode durations with Delta infections in index cases observed could also be due to repeat infections in some index cases, which have been shown to result in shorter episode durations.[27] Episode duration based on Ct<30 (as a proxy for higher viral load) was longer in females. This association of longer episode duration in females has not been observed in other studies, and warrants further investigation.

Very limited data are available on SARS-CoV-2 serial interval, especially so for people LWH. Previous estimates have ranged between 4 to 7.5 days[2, 19] which is similar to our estimate of 6 days. We did not find any index characteristics related to longer serial intervals, but longer serial intervals were observed in contacts older than 35 years, and those LWH. Individuals with compromised immune systems (PLWH, elderly) may still be able to be infected with SARS-CoV-2 towards the end of the index episode when viral loads are lower.

From our results, there were fewer secondary cases in contacts who smoked. While evidence exists that smoking may lead to poorer COVID-19 outcomes,[29] other studies have also found a negative association with smoking and SARS-CoV-2 infection.[29, 30] The latter association may be a result of confounding biases including socio-economic status and healthcare seeking behaviour,[29] or possible decreased test sensitivity[30] or could reflect a true biological interaction.

Our study had limitations. We assumed the first household member presenting with symptoms was the index case. If the true index cases were asymptomatic, we would have underestimated the serial interval, although HCIR estimates should not be greatly affected. By excluding individuals seropositive at baseline with no SARS-CoV-2 infection during follow-up, we assumed a 100% protection from previous infections, which is likely not correct, and in turn may have overestimated the HCIR. When including these individuals, the HCIR reduced by 8%. We were unable to reach the planned sample size for contacts of index cases LWH, and may have been underpowered to detect some differences, especially when stratifying by immunocompromised status.

In conclusion, in two communities in South African households, HCIR was higher than in previous studies[14] but was not influenced by HIV status. Episode duration at high viral loads and serial interval was increased for people LWH. Although HIV may not be the primary driver in SARS-CoV-2 transmission, it may still play a role, especially if PLWH are not virally suppressed. The SARS-CoV-2 pandemic impacted several health programs, including HIV testing and care. During initial lockdowns there was a decline in HIV testing and antiretroviral therapy (ART) initiations, which gradually returned to pre-lockdown levels after the lockdown in South Africa[31] and other Sub-Saharan African countries.[32] Sustaining and strengthening HIV treatment and care programs should be a focus moving forward.

## Data Availability

The investigators welcome enquiries about possible collaborations and requests for access to the dataset. Data will be shared after approval of a proposal and with a signed data access agreement. Investigators interested in more details about this study, or in accessing these resources, should contact the corresponding author.

## Funding

This research was funded by the Wellcome Trust (Grant number 221003/Z/20/Z) in collaboration with the Foreign, Commonwealth and Development Office, United Kingdom. For the purpose of open access, the author has applied a CC BY-ND public copyright licence to any Author Accepted Manuscript version arising from this submission.

## Acknowledgements

All individuals participating in the study, field teams for their hard work and dedication to the study, the CRDM laboratory team, Andrew Whitelaw, June Fabian, Themba Radebe and Thoko Sifunda from the safety committee.

This study protocol was adapted from the World Health Organization Unity Studies Early Investigation Protocol for Household transmission investigation protocol for COVID-19 infection.

## Potential conflicts of interest

CC has received grant support from Sanofi Pasteur, US CDC, Welcome Trust, Programme for Applied Technologies in Health (PATH), Bill & Melinda Gates Foundation and South African Medical Research Council (SA-MRC). Nicole Wolter and Anne von Gottberg report receiving grant funds from Sanofi and Gates Foundation. All other authors have no competing/conflict of interest.

## Author contributions

Conception and design of study: JK, SW, NAM, AvG, JNB, MW, LL, ST, CC

Data collection and laboratory processing: AvG, JNB, DGA, AB, KN, NW, LG, LN, JC, LdG, RK

Analysis and interpretation: JK, SW, ST, CC

Accessed and verified underlying data: JK, MN, LG, LM, JC

Drafted the Article: JK

All authors critically reviewed the Article. All authors had access to all the data reported in the study.

