## Supplement for "Household transmission of SARS-CoV-2 from adult index cases living with and without HIV in South Africa, 2020-2021: a case-ascertained, prospective observational household transmission study"

### Supplementary methods

*Study population*

We conducted the study at two sites, Klerksdorp and Soweto. Klerksdorp is located in the municipality of Matlosana in North West Province and has a population of over 385,000 people and is 115 km$^2$. The households of study participants in Klerksdorp include mostly single-family houses and shacks. Prevalence of HIV in the North West Province was estimated at 18% in 2020 for individuals >15 years.[1] Soweto is a township outside of Johannesburg, located in the Gauteng Province where the HIV prevalence for individuals >15 years was estimated at 15% in 2020.[1] Its population is 1.5 million and encompasses 150 km$^2$. Housing structures include, but are not limited to single-family houses, multi-unit dwellings, shacks within informal settlements and in the backyards of formal residences, and hostels. Clinic catchment areas were defined as Kenneth Kaunda District in North West Province and sub-district Region D of City of Johannesburg district in Gauteng Province),

*Sample size*

We aimed to assess a significant difference in the household cumulative infection risk (HCIR) between household contacts exposed to SARS-CoV-2 by a HIV-infected vs HIV-uninfected index case for a 95% confidence interval and 80% power. We assumed an average household size of 5 individuals, a HIV prevalence among index cases presenting with COVID-19-like-illness of 40% (based on HIV prevalence for influenza-like illness),[2] and a HCIR of 10% in household members exposed to an HIV-uninfected index case and a HCIR of 20% among household members exposed to a HIV-infected index case. The resulting total sample size for this study was 440 exposed household members: 264 exposed household members from households with an HIV-uninfected index case and 176 exposed household members from households with an index case LWH.

*Screening for index cases*

Screening for index cases started in October 2020 and continued September 2021, and was done at three clinics in Klerksdorp (Jouberton, Alabama and Grace Mokhomo clinics), and five in Soweto (Lillian Ngoyi, Zola, Mofolo and Chaiwelo Community Health Centres; and Mofolo clinic). Screening started in October 2020 and continued until June 2021 in Klerksdorp and September 2021 in Soweto. In July 2021, the Soweto site was reduced to two clinics.

Study staff screened clinic attendees 18 years and older on week days for suspected COVID-19 including any of the following symptoms: fever, cough, sore throat, difficulty breathing, myalgia, headaches, running nose and diarrhoea; with symptom onset ≤5 days prior to screening. Two nasopharyngeal swabs were collected from consenting eligible individuals; one of which was transported in viral or universal transport medium to the National Institute for Communicable Diseases (NICD) for SARS-CoV-2 RNA detection using real-time reverse transcription polymerase chain reaction (rRT-PCR), and the other was used for point-of-care SARS-CoV-2 testing using the rapid antigen detection kit (implemented February 2021, Standard Q COVID-19 Ag test, SD Biosensor, Chungcheongbuk-do, South Korea). All positive antigen tests were verified with rRT-PCR. Staff also completed a case investigation form to collect demographic information, symptoms, and underlying medical conditions, including HIV infection. In addition to active screening at clinics, individuals who tested positive for SARS-CoV-2 meeting study case definitions were also referred from the influenza-like illness surveillance program in Klerksdorp[2] and by contact tracing teams in the site districts as part of the national public health response to COVID-19 in South Africa. Referred individuals were included if a documented SARS-CoV-2 positive test was available.

*Household enrolment*

We approached households of individuals who tested positive for SARS-CoV-2, with symptom onset less than 7 days prior (up to 5 days prior to screening with another 48 hours for testing to be completed), and who reported at least two additional household members. We excluded households who resided outside the clinics' catchment areas, if household contacts reported symptoms in the 14 days prior to screening, or if the index cases were hospitalised immediately following screening). Once the head of household provided permission, all household members were approached for consenting and enrolment. Households with ≥3 eligible members, of whom ≥70% consented to participate were enrolled. During enrolment we collected information on household characteristics such as household size, caretaking roles, sleeping arrangements and housing; and demographics, underlying medical conditions, education, employment, smoking, alcohol use and vaccination information. Households that withdrew within 10 days from index symptom start date were excluded from the analysis.

*HIV testing*

Point of care rapid HIV enzyme-linked immunosorbent assay (ELISA) testing (WANTAI HIV 1+2Ab Rapid test, Beijing Wantai Biological, Beijing, China) was offered to all individuals at screening with an unknown HIV status, or for whom a documented negative HIV result was from > 6 months previously. For consenting household contacts HIV status was determined using rapid HIV tests if unknown or a documented negative status was not available within the previous 6 months. For children aged 10 years and younger whose mother had a documented HIV negative status during pregnancy, HIV status was considered negative. Where participants did not agree to rapid testing at point of care, but did consent to HIV testing, residual serum was tested for HIV antibodies using an ELISA assay at NICD. Data on recent CD4+ T cell count and HIV viral load was collected for individuals living with HIV. If these data were not available, samples were collected and viral load tested by quantitative PCR (Roche Cobas Ampliprep/Cobas Taqman HIV-1 test, Roche Diagnostics, Mannheim, German) at the NICD.

*SARS-CoV-2 detection*

Nasopharyngeal (screening) and nasal (follow-up) specimens were collected using nylon flocked swabs and transported in viral or universal transport medium to the NICD for testing. Total nucleic acids were extracted from 200µl of sample using a MagNA Pure 96 automated extractor and the DNA/Viral NA Small Volume v2.0 extraction kit (Roche Diagnostics, Mannheim, Germany). The E, RdRp and N SARS-CoV-2 genes were detected by rRT-PCR, using the Allplex™ 2019-nCoV kit

(Seegene Inc., Seoul, South Korea). Specimens were considered positive for SARS-CoV-2 if the cycle threshold ($C_t$) value was below 40 for any of the gene targets.

*SARS-CoV-2 variants*

The first SARS-CoV-2 positive specimen for each participant was subjected to the Allplex™ SARS-CoV-2 Variants I and II PCR assays (Seegene Inc., Seoul, Korea). The Variants I assay targets the RdRp gene, HV69/70 deletion, N501Y and E484K mutations; and the Variants II assay targets the L452R, W152C, K417T and K417N mutations; allowing differentiation between the Alpha (B.1.1.7 Pango lineage), Beta (B.1.351), Delta (B.1.617.2), Gamma (P.1) variants of concern, and the US-California variant of interest (B.1.429).

In addition, the same specimen was sequenced on the Ion Torrent Genexus platform using AmpliSeq for SARS-CoV-2 (Illumina), which is based on the ARTIC SARS-CoV-2 sequencing protocol (https://www.protocols.io/view/illumina-nextera-dna-flex-library-construction-and-bhjgj4jw). Genomes were assembled using the Exatype SARS-CoV-2 pipeline (https://sars-cov-2.exatype.com/), which includes de-duplicating sequenced reads data, base-calling, de-multiplexing, removal of amplicon primer sequences prior to variant calling, mapping to a reference using Examap and finally consensus sequence generation. These consensus sequences were manually inspected and polished using Aliview v1.27 (http://ormbunkar.se/aliview/). The reference genome used throughout the assembly process was NC_045512.2 (Accession number: MN908947.3). Clade and lineage assignments were made using the online Nextclade (https://clades.nextstrain.org/) and PANGOLin (https://pangolin.cog-uk.io/) applications, which also enable identification of known variants of concern as well as novel mutations. Only specimens with a Ct value <35 on at least one gene target in the initial rRT-PCR was sequenced. If the first specimen in the episode was not successfully characterised (due to high Ct values in initial rRT-PCR or poor sample quality), a second specimen was characterised by variant PCR and sequencing.

Variants were assigned firstly based on sequencing results where sequence coverage/reads was sufficient for clade and lineage classification, followed by the variant PCR results when sequence coverage/reads was low, and then based on variants detected within the household if no sequencing or variant PCR result was available. Where no variant-specific mutations were detected in the PCR, and the RdRp gene Ct value was <35, and internal controls for both assays were detected, the episodes were classified as attributed to a non-Alpha/Beta/Delta variant. Where more than one variant was detected within one SARS-CoV-2 cluster, specimens were retested from fresh aliquots of the same sample to confirm results. Confirmed mixed variant clusters were excluded from HCIR analyses.

*Serology*

We used an ELISA to detect antibodies against SARS-CoV-2 spike protein as previously described[3]. In summary, recombinant trimeric spike protein was coated at 2 µg/ml onto 96-well, high-binding plates and incubated overnight at 4°C. The plates were washed and incubated at 37 °C for 1–2 hours in blocking buffer (5% skimmed milk powder, 0.05% Tween 20, 1× PBS). Serum samples were added at 1:100 dilution, following a one hour incubation at 37 °C and anti-human horseradish peroxidase-conjugated antibody was added for another one hour at 37°C. The signal was developed with OneStep TMB substrate (Thermo Fisher Scientific, USA) for 5 min at room temperature, followed by addition of 1 M $H_2SO_4$ stop solution. Absorbance at 450 nm was measured and specimens with optical density (OD)>0.4 were considered positive for anti-spike antibodies. For the nucleocapsid protein we performed the assay on the Cobas e601 instrument (Roche Diagnostics), and we considered a cutoff index (COI) >1.0 as positive for anti-nucleocapsid antibodies.

### Supplementary results

*Serology*

At baseline, 67% (83/123) of index cases for whom serology results were available were already seropositive for SARS-CoV-2, which increased to 98% (107/109) after six weeks of follow-up. Two individuals had a rRT-PCR-confirmed SARS-CoV-2 episode, but were still seronegative for SARS-CoV-2 antibodies at the end of follow-up. The first individual who did not seroconvert was a HIV-uninfected male. Episode duration was 6 days, with only one positive swab (collected at screening) with a minimum Ct value of 38. There was no secondary transmission (0/2) detected in the household linked to the illness episode. The second was a female living with HIV, with an episode duration of 9 days and minimum Ct value of 26. There was an 80% (4/5) attack rate in the household. No HIV viral load or CD4+ T cell count were available.

Paired baseline and exit sera were available in 72% (331/457) of contacts. In those with paired sera, 39% (128/331) where already seropositive at baseline, 57% (73/128) of whom had a PCR-confirmed SARS-CoV-2 infection during follow-up. Of those initially seronegative, 47% (95/203) were seropositive after six weeks of follow-up; 80% (76/95) of whom had at least one respiratory specimen positive for SARS-CoV-2 positive specimen. Twenty percent (19/95) of contacts initially seronegative who had at least one respiratory specimen positive for SARS-CoV-2 positive did not seroconvert by the end of the follow-up period. Of the 91 index cases and 331 contacts with paired sera, no sero-reversions were detected.

*Mixed SARS-CoV-2 variant clusters*

We identified four households where multiple SARS-CoV-2 variants were detected within one cluster. These were two Beta and Delta clusters, one Alpha and Beta cluster, and one Beta and C.1.2 cluster (Supplementary Figure 3).

*Limitations*

Although we reached our overall sample size, we only managed to enrol 103 exposed household members from households with an index case LWH (59% of target). The HIV prevalence amongst screened patients were lower than anticipated (9% vs expected 40% as seen in surveillance for influenza-like illness). Furthermore, very few index cases were immunosuppressed, likely due to a high proportion of PLWH being in care. Individuals who are not on ART may be less likely to seek care and be identified as SARS-CoV-2 index cases.

### Supplementary tables

**Supplementary Table 1.** Factors associated with SARS-CoV-2 household transmission in index cases and acquisition in household contacts, Klerksdorp and Soweto, South Africa, 2020-2021, (n=373).

| Characteristic | HCIR*, n/N (%) | Univariate, OR (95%CI) | Multivariable, aOR (95%CI) |
|---|---|---|---|
| **Index characteristics** | | | |
| **Site** | | | |
| Klerksdorp | 95/174 (54.6) | Reference | |
| Soweto | 125/199 (62.8) | 1.4 (0.7-2.8) | 1.1 (0.5-2.1) |
| **Age (years)** | | | |
| 18-34 | 54/108 (50.0) | Reference | Reference |
| 35-59 | 137/218 (62.8) | 1.9 (0.9-4.2) | **3.4 (1.5-7.8)** |
| ≥60 | 29/47 (61.7) | 1.8 (0.6-6.0) | **3.1 (1.0-10.1)** |
| **Sex** | | | |
| Male | 64/111 (57.7) | 0.9 (0.4-1.9) | |
| Female | 156/262 (59.5) | Reference | |
| **HIV** | | | |
| HIV-infected | 51/85 (60.0) | 1.0 (0.4-2.3) | |
| HIV-uninfected | 163/279 (58.4) | Reference | |
| Unknown HIV-status | 6/9 (66.7) | 0.8 (0.1-11.4) | |
| **CD4+ T cell count** | | | |
| HIV-infected, not immune suppressed† | 30/52 (57.7) | Reference | Reference |
| HIV-infected, immune suppressed† | 8/13 (61.5) | 1.6 (0.2-13.5) | 2.5 (0.4-15.3) |
| HIV-infected, no CD4+ T cell count | 13/20 (65.0) | 1.5 (0.2-8.7) | 1.2 (0.2-5.8) |
| HIV-uninfected | 163/279 (58.4) | 1.2 (0.4-3.3) | 1.8 (0.7-4.9) |
| Unknown HIV-status | 6/9 (66.7) | 1.0 (0.1-15.3) | 0.5 (0.0-5.5) |
| **Underlying illness‡** | | | |
| No | 172/293 (58.7) | 0.8 (0.3-1.8) | |
| Yes | 48/75 (64.0) | Reference | |
| Unknown | 0/5 (0.0) | Excluded | |
| **Body-mass index** | | | |
| Underweight | 3/9 (33.3) | 0.2 (0.0-2.2) | **0.1 (0.0-0.8)** |
| Normal weight | 68/108 (63.0) | Reference | Reference |
| Overweight | 44/67 (65.7) | 1.1 (0.4-3.2) | 0.6 (0.2-1.5) |
| Obese | 105/184 (57.1) | 0.8 (0.4-1.7) | **0.4 (0.2-0.9)** |
| Unknown | 0/5 (0.0) | Excluded | Excluded |
| **Crowding** | | | |
| No | 56/88 (63.6) | 1.5 (0.7-3.3) | |
| Yes | 164/285 (57.5) | Reference | |
| **Child <5 years in home** | | | |
| No | 190/316 (60.1) | 1.8 (0.7-5.0) | |
| Yes | 30/57 (52.6) | Reference | |
| **Current cigarette smoke (self-reported)** | | | |
| No | 203/328 (61.9) | 2.8 (1.0-8.0) | |
| Yes | 17/40 (42.5) | Reference | |
| Unknown | 0/5 (0.0) | Excluded | |
| **Episode duration** | | | |

| Characteristic | HCIR*, n/N (%) | Univariate, OR (95%CI) | Multivariable, aOR (95%CI) |
|---|---|---|---|
| 1-10 days | 64/122 (52.5) | Reference | |
| 11-21 days | 53/95 (55.8) | 1.5 (0.6-3.6) | |
| >21 days | 101/151 (66.9) | **2.3 (1.0-5.3)** | |
| Positive at last visit | 2/5 (40.0) | 0.5 (0.0-8.7) | |
| **Fever** | | | |
| No | 110/187 (58.8) | Reference | |
| Yes | 110/186 (59.1) | 1.1 (0.5-2.2) | |
| **Minimum Ct value** | | | |
| >35 | 9/39 (23.1) | Reference | Reference |
| 25-35 | 86/139 (61.9) | **8.3 (2.4-28.1)** | **7.5 (2.2-26.0)** |
| <25 | 123/192 (64.1) | **8.7 (2.7-28.4)** | **5.3 (1.6-17.6)** |
| Unknown | 2/3 (66.7) | 10.2 (0.3-402.1) | 11.3 (0.4-355.6) |
| **Serostatus at follow-up end** | | | |
| Seronegative | 4/7 (57.1) | 0.5 (0.0-7.4) | |
| Seropositive | 182/293 (62.1) | Reference | |
| Unknown | 34/73 (46.6) | 0.4 (0.2-1.1) | |
| **SARS-CoV-2 variant** | | | |
| non-Alpha/Beta/Delta | 18/28 (64.3) | 1.3 (0.4-4.2) | 2.1 (0.7-6.3) |
| Alpha | 16/20 (80.0) | 3.3 (0.7-14.8) | 3.4 (0.8-15.6) |
| Beta | 144/253 (56.9) | Reference | Reference |
| Delta | 40/49 (81.6) | **3.6 (1.2-10.7)** | **4.6 (1.5-14.4)** |
| Unknown | 2/23 (8.7) | 0.0 (0.0-0.3) | 0.1 (0.0-0.5) |
| **Contact characteristics** | | | |
| **Age (years)** | | | |
| <5 | 5/19 (26.3) | Reference | Reference |
| 5-12 | 40/73 (54.8) | 4.2 (1.0-17.6) | 1.9 (0.4-7.9) |
| 13-17 | 43/60 (71.7) | **15.8 (3.4-74.0)** | **7.1 (1.5-33.9)** |
| 18-34 | 56/95 (59.0) | **6.4 (1.6-26.1)** | **4.4 (1.0-18.4)** |
| 35-59 | 56/90 (62.2) | **6.3 (1.5-26.4)** | 4.4 (1.0-18.6) |
| ≥60 | 20/36 (55.6) | **6.1 (1.2-30.2)** | 2.9 (0.6-14.4) |
| **Sex** | | | |
| Male | 83/160 (51.9) | Reference | |
| Female | 137/213 (64.3) | **2.0 (1.2-3.4)** | |
| **HIV** | | | |
| HIV-uninfected | 82/147 (55.8) | 1.1 (0.4-3.1) | |
| HIV-infected | 16/28 (57.1) | Reference | |
| HIV status unknown | 122/198 (61.6) | 1.3 (0.5-3.5) | |
| **Underlying illness‡** | | | |
| No | 189/323 (58.5) | Reference | |
| Yes | 30/42 (71.4) | 2.6 (1.0-6.8) | |
| Unknown | 1/8 (12.5) | 0.1 (0.0-0.8) | |
| **Body-mass index** | | | |
| Underweight | 15/25 (60.0) | 1.0 (0.3-3.0) | |
| Normal weight | 96/173 (55.5) | Reference | |
| Overweight | 44/77 (57.1) | 1.2 (0.6-2.4) | |
| Obese | 64/90 (71.1) | **2.4 (1.2-4.9)** | |
| Unknown | 1/8 (12.5) | 0.1 (0.0-1.0) | |

| Characteristic | HCIR*, n/N (%) | Univariate, OR (95%CI) | Multivariable, aOR (95%CI) |
|---|---|---|---|
| **Current cigarette smoke (self-reported)** | | | |
| No | 197/320 (61.6) | **2.8 (1.2-6.2)** | **3.9 (1.7-9.0)** |
| Yes | 22/49 (44.9) | Reference | Reference |
| Unknown | 1/4 (25.0) | 0.4 (0.0-10.4) | Excluded |
| **Sleep in same room as index** | | | |
| No | 150/261 (57.5) | Reference | |
| Yes | 70/112 (62.5) | 1.2 (0.7-2.1) | |
| **Index is main caregiver** | | | |
| No | 183/313 (58.5) | 1.2 (0.6-2.6) | |
| Yes | 37/60 (61.7) | Reference | |

OR – Odds ratio, aOR – Adjusted odds ratio. *Household cumulative infection risk (HCIR) excludes household members seropositive at baseline with no SARS-CoV-2 detected on PCR during follow-up and households with mixed SARS-CoV-2 variant clusters. †Immune suppressed defined as CD4+ T cell count <200 cells/ml. ‡Underlying illness: self-reported history of diabetes, hypertension, asthma, lung disease, heart disease, stroke, spinal cord injury, epilepsy, cancer, liver disease, renal disease, pre-maturity. Bold face font denotes $p<0.05$.

**Supplementary Table 2.** Factors associated with SARS-CoV-2 episode duration (irrespective of Ct value) in index cases, Klerksdorp and Soweto, South Africa, 2020-2021, (n=123). HR – Hazard ratio*

| Characteristic | Episode duration, days, Mean ± SD (Range) | Univariate, HR* (95%CI) | Multivariable, aHR (95%CI) |
|---|---|---|---|
| **Site** | | | |
| Klerksdorp | 17.0±11.6 (4.0-45.0) | Reference | Reference |
| Soweto | 21.2±12.6 (3.0-45.0) | **0.6 (0.5-0.9)** | **0.4 (0.3-0.7)** |
| **Age (years)** | | | |
| 18-34 | 16.5±11.3 (4.0-41.0) | Reference | Reference |
| 35-59 | 18.6±12.6 (3.0-45.0) | 0.7 (0.5-1.1) | **0.4 (0.2-0.6)** |
| ≥60 | 27.3±10.0 (6.0-43.0) | **0.5 (0.3-0.8)** | **0.2 (0.1-0.5)** |
| **Sex** | | | |
| Male | 18.5±13.2 (3.0-45.0) | Reference | Reference |
| Female | 19.3±11.9 (3.0-45.0) | 1.1 (0.7-1.6) | 0.8 (0.5-0.0) |
| **HIV** | | | |
| HIV-infected | 17.2±12.0 (3.0-45.0) | 0.8 (0.5-1.2) | 1.5 (0.8-2.7) |
| HIV-uninfected | 19.6±12.4 (3.0-45.0) | Reference | Reference |
| Unknown HIV-status | 18.5±10.6 (11.0-26.0) | 1.3 (0.3-5.6) | 0.9 (0.2-4.5) |
| **CD4+ T cell count** | | | |
| HIV-infected, not immune suppressed† | 18.8±12.8 (3.0-45.0) | Reference | Reference |
| HIV-infected, immune suppressed† | 14.1±9.0 (7.0-33.0) | 1.4 (0.5-4.3) | 1.0 (0.3-3.7) |
| HIV-infected, no CD4+ T cell count | 14.1±9.0 (7.0-33.0) | 1.9 (0.8-4.7) | 0.9 (0.3-2.5) |
| HIV-uninfected | 19.6±12.4 (3.0-45.0) | 1.0 (0.6-1.6) | Omitted |
| Unknown HIV-status | 18.5±10.6 (11.0-26.0) | 1.4 (0.3-6.2) | Omitted |
| **Underlying illness‡** | | | |
| No | 19.1±12.4 (3.0-45.0) | Reference | |
| Yes | 19.8±12.2 (3.0-41.0) | 0.8 (0.5-1.3) | |
| Unknown | 9.0±0.0 (9.0-9.0) | 4.0 (1.0-17.0) | |
| **Body-mass index** | | | |
| Underweight | 13.0±4.6 (9.0-18.0) | **3.0 (0.9-10.1)** | **5.8 (1.5-22.1)** |
| Normal weight | 20.1±12.9 (6.0-45.0) | Reference | Reference |
| Overweight | 19.9±14.5 (3.0-43.0) | 1.0 (0.6-1.7) | 1.7 (0.9-3.2) |
| Obese | 18.8±11.3 (4.0-44.0) | 1.2 (0.8-1.9) | **2.0 (1.2-3.5)** |
| Unknown | 9.0±0.0 (9.0-9.0) | 4.6 (1.1-19.9) | 2.8 (0.6-14.2) |
| **Crowding** | | | |
| No | 18.0±11.7 (3.0-43.0) | Reference | |
| Yes | 19.6±12.5 (3.0-45.0) | 0.8 (0.5-1.2) | |
| **Child <5 years in home** | | | |
| No | 19.5±12.2 (3.0-45.0) | 0.7 (0.4-1.3) | |
| Yes | 16.6±12.5 (4.0-35.0) | Reference | |
| **Current cigarette smoke (self-reported)** | | | |
| No | 19.9±12.5 (3.0-45.0) | 0.6 (0.3-1.1) | |
| Yes | 15.3±10.3 (3.0-37.0) | Reference | |
| Unknown | 9.0±0.0 (9.0-9.0) | 2.8 (0.6-12.5) | |
| **Fever** | | | |
| No | 15.8±10.5 (3.0-45.0) | Reference | |
| Yes | 22.4±13.1 (3.0-45.0) | **0.7 (0.5-0.9)** | |
| **Minimum Ct value** | | | |
| >35 | 10.8±7.0 (3.0-26.0) | **0.4 (0.2-0.8)** | 1.8 (0.9-3.6) |

| Characteristic | Episode duration, days, Mean ± SD (Range) | Univariate, HR* (95%CI) | Multivariable, aHR (95%CI) |
|---|---|---|---|
| 25-35 | 15.2±11.8 (3.0-45.0) | Reference | Reference |
| <25 | 24.1±11.5 (9.0-43.0) | **0.3 (0.1-0.5)** | **0.5 (0.3-0.9)** |
| Unknown | 8.0±0 (8.0-8.0) | 2.2 (0.3-17.0) | 12.2 (1.4-103.2) |
| **Serostatus at follow-up end** | | | |
| Seronegative | 7.5±2.1 (6.0-9.0) | Reference | Reference |
| Seropositive | 20.6±12.4 (3.0-45.0) | **0.1 (0.0-0.5)** | **0.0 (0.0-0.3)** |
| Unknown | 13.5±10.2 (3.0-38.0) | 0.2 (0.1-1.0) | 0.1 (0.0-0.7) |
| **SARS-CoV-2 variant** | | | |
| non-Alpha/Beta/Delta | 13.6±9.8 (4.0-38.0) | 2.4 (1.3-4.6) | **4.0 (1.9-8.2)** |
| Alpha | 21.4±15.1 (7.0-41.0) | 0.9 (0.4-1.9) | 0.8 (0.3-2.1) |
| Beta | 20.4±12.9 (3.0-45.0) | Reference | Reference |
| Delta | 20.1±9.9 (4.0-41.0) | 1.2 (0.7-2.1) | **2.6 (1.4-5.0)** |
| Unknown | 11.4±7.0 (5.0-28.0) | 3.0 (1.5-5.9) | 1.6 (0.7-3.6) |

*Estimated using Weibull accelerated failure time regression adjusted for clustering by site and household. Hazard ratio <1 corresponds to longer episode duration than reference group used. †Immune suppressed defined as CD4+ T cell count <200 cells/ml. ‡Underlying illness: self-reported history of diabetes, hypertension, asthma, lung disease, heart disease, stroke, spinal cord injury, epilepsy, cancer, liver disease, renal disease, pre-maturity. Bold face font denotes p<0.05.

**Supplementary Table 3.** Factors associated with SARS-CoV-2 episode duration (Ct value<30) in index cases, Klerksdorp and Soweto, South Africa, 2020-2021, (n=123). HR – Hazard ratio*

| Characteristic | Episode duration, days, Mean ± SD (Range) | Univariate, HR* (95%CI) | Multivariable, aHR (95%CI) |
|---|---|---|---|
| **Site** | | | |
| Klerksdorp | 6.9±2.2 (3.0-13.0) | Reference | Reference |
| Soweto | 7.0±2.6 (2.0-17.0) | 0.6 (0.4-1.0) | 0.7 (0.4-1.2) |
| **Age (years)** | | | |
| 18-34 | 6.6±2.8 (3.0-17.0) | Reference | Reference |
| 35-59 | 7.0±2.2 (2.0-13.0) | 1.0 (0.6-1.6) | 1.0 (0.5-1.8) |
| ≥60 | 7.3±2.5 (4.0-12.0) | **0.3 (0.1-0.6)** | 0.6 (0.2-1.5) |
| **Sex** | | | |
| Male | 6.7±2.4 (3.0-12.0) | Reference | Reference |
| Female | 7.0±2.5 (2.0-17.0) | 0.7 (0.4-1.1) | **0.5 (0.3-0.9)** |
| **HIV** | | | |
| HIV-infected | 7.3±2.8 (2.0-12.0) | **0.4 (0.2-0.7)** | **0.4 (0.1-0.9)** |
| HIV-uninfected | 6.8±2.3 (3.0-17.0) | Reference | Reference |
| Unknown HIV-status | 9.0±0 (9.0-9.0) | 0.6 (0.1-4.4) | 0.6 (0.1-4.9) |
| **CD4+ T cell count** | | | |
| HIV-infected, not immune suppressed† | 7.2±3.1 (2.0-12.0) | Reference | Reference |
| HIV-infected, immune suppressed† | 7.3±2.1 (5.0-9.0) | **4.1 (1.1-10.9)** | 2.3 (0.5-10.3) |
| HIV-infected, no CD4+ T cell count | 7.5±3.3 (4.0-12.0) | 3.3 (1.0-9.3) | 0.6 (0.1-3.0) |
| HIV-uninfected | 6.8±2.3 (3.0-17.0) | **4.4 (2.1-21.6)** | Omitted |
| Unknown HIV-status | 9.0±0 (9.0-9.0) | 2.7 (0.3-0.0) | Omitted |
| **Underlying illness‡** | | | |
| No | 7.0±2.6 (2.0-17.0) | Reference | |
| Yes | 6.6±2.0 (3.0-11.0) | 1.5 (0.9-2.5) | |
| Unknown | 9.0±0 (9.0-9.0) | 0.7 (0.1-5.5) | |
| **Body-mass index** | | | |
| Underweight | 10.5±2.1 (9.0-12.0) | 0.8 (0.2-3.6) | 0.6 (0.1-2.7) |
| Normal weight | 6.7±1.7 (4.0-10.0) | Reference | Reference |
| Overweight | 6.1±2.2 (3.0-12.0) | **2.7 (1.3-5.3)** | 2.1 (0.9-5.0) |
| Obese | 7.2±2.7 (2.0-17.0) | 1.7 (1.0-3.0) | 1.3 (0.6-2.7) |
| Unknown | 9.0±0 (9.0-9.0) | 1.2 (0.2-9.4) | 0.6 (0.1-5.2) |
| **Crowding** | | | |
| No | 6.6±2.2 (3.0-13.0) | Reference | |
| Yes | 7.1±2.5 (2.0-17.0) | 0.7 (0.4-1.0) | |
| **Child <5 years in home** | | | |
| No | 6.9±2.5 (2.0-17.0) | 0.8 (0.4-1.6) | |
| Yes | 6.8±1.2 (5.0-9.0) | Reference | |
| **Current cigarette smoke (self-reported)** | | | |
| No | 6.8±2.3 (2.0-13.0) | 1.2 (0.6-2.3) | |
| Yes | 7.8±3.6 (5.0-17.0) | Reference | |
| Unknown | 9.0±0 (9.0-9.0) | 0.8 (0.1-6.5) | |
| **Fever** | | | |
| No | 6.3±2.4 (2.0-13.0) | Reference | |
| Yes | 7.4±2.4 (3.0-17.0) | **1.1 (0.7-1.7)** | |
| **Minimum Ct value** | | | |

| Characteristic | Episode duration, days, Mean ± SD (Range) | Univariate, HR* (95%CI) | Multivariable, aHR (95%CI) |
|---|---|---|---|
| >35 | No observations | No observations | No observations |
| 25-35 | 11.1±6.5 (3.0-31.0) | Reference | Reference |
| <25 | 24.1±11.5 (9.0-43.0) | **0.7 (0.4-1.2)** | 1.1 (0.6-2.1) |
| Unknown | No observations | No observations | No observations |
| **Serostatus at follow-up end** | | | |
| Seronegative | 4.0±0 (4.0-4.0) | Reference | Reference |
| Seropositive | 7.0±2.5 (2.0-17.0) | **0.2 (0.0-1.2)** | **0.01 (0.01-0.2)** |
| Unknown | 6.6±2.1 (3.0-10.0) | 0.2 (0.0-1.9) | **0.02 (0.01-0.3)** |
| **SARS-CoV-2 variant** | | | |
| non-Alpha/Beta/Delta | 6.8±1.6 (5.0-10.0) | 1.3 (0.7-2.7) | 1.7 (0.8-3.7) |
| Alpha | 6.0±2.2 (3.0-9.0) | 1.6 (0.7-3.8) | 1.6 (0.6-4.2) |
| Beta | 7.2±2.6 (3.0-17.0) | Reference | Reference |
| Delta | 6.4±2.1 (3.0-11.0) | **0.4 (0.2-0.8)** | 1.1 (0.5-2.5) |
| Unknown | 6.0±5.7 (2.0-10.0) | 1.1 (0.3-4.5) | 1.0 (0.2-4.5) |

*Estimated using Weibull accelerated failure time regression adjusted for clustering by site and household. Hazard ratio <1 corresponds to longer episode duration than reference group used. †Immune suppressed defined as CD4+ T cell count <200 cells/ml. ‡Underlying illness: self-reported history of diabetes, hypertension, asthma, lung disease, heart disease, stroke, spinal cord injury, epilepsy, cancer, liver disease, renal disease, pre-maturity. Omitted categories of co-variates were co-linear in multivariable model. Bold face font denotes p<0.05.

**Supplementary Table 4.** Factors associated with serial interval of SARS-CoV-2 in cases with symptomatic illness, Klerksdorp and Soweto, South Africa, 2020-2021, (n=62). HR – Hazard ratio*

| Characteristic | Serial interval, days, Mean ± SD (Range) | Univariate, HR* (95%CI) | Multivariable, aHR (95%CI) |
|---|---|---|---|
| **Index characteristics** | | | |
| **Site** | | | |
| Klerksdorp | 5.8±4.1 (1.0-21.0) | Reference | |
| Soweto | 7.5±4.0 (2.0-20.0) | 0.3 (0.1-1.3) | 0.3 (0.1-1.3) |
| **Age (years)** | | | |
| 18-34 | 6.6±4.6 (1.0-15.0) | Reference | |
| 35-59 | 6.4±3.6 (2.0-21.0) | 0.6 (0.1-3.0) | |
| ≥60 | 7.0±6.4 (2.0-20.0) | 0.9 (0.1-7.9) | |
| **Sex** | | | |
| Male | 6.7±4.6 (2.0-21.0) | 0.9 (0.2-3.9) | |
| Female | 6.5±3.9 (1.0-20.0) | Reference | |
| **HIV** | | | |
| HIV-infected | 9.3±7.9 (2.0-21.0) | 0.2 (0.0-1.4) | |
| HIV-uninfected | 6.1±3.2 (1.0-15.0) | Reference | |
| **Underlying illness‡** | | | |
| No | 6.9±4.4 (1.0-21.0) | Reference | |
| Yes | 17.0±5.5 (2.0-14.0) | 1.5 (0.4-6.1) | |
| **Body-mass index** | | | |
| Normal weight | 7.2±5.8 (2.0-21.0) | 0.7 (0.1-4.8) | |
| Overweight | 6.3±3.7 (2.0-15.0) | Reference | |
| Obese | 6.4±3.7 (1.0-20.0) | 0.7 (0.1-3.5) | |
| **Crowding** | | | |
| No | 5.9±4.1 (1.0-15.0) | Reference | |
| Yes | 6.8±4.1 (2.0-21.0) | 0.8 (0.2-3.3) | |
| **Child <5 years in home** | | | |
| No | 6.5±4.2 (1.0-21.0) | 0.4 (0.0-3.5) | |
| Yes | 6.9±3.6 (2.0-12.0) | Reference | |
| **Current cigarette smoke (self-reported)** | | | |
| No | 6.7±4.2 (1.0-21.0) | 0.2 (0.0-5.9) | |
| Yes | 4.3±0.5 (4.0-5.0) | Reference | |
| **Episode duration** | | | |
| 1-10 days | 6.8±4.0 (2.0-15.0) | 2.0 (0.3-12.3) | |
| 11-21 days | 7.4±4.6 (1.0-21.0) | Reference | |
| >21 days | 5.9±4.0 (2.0-20.0) | 3.9 (0.6-24.0) | |
| Unknown | | | |
| **Fever** | | | |
| No | 7.0±4.7 (1.0-20.0) | 1.0 (0.3-4.0) | |
| Yes | 6.2±3.7 (2.0-21.0) | Reference | |
| **Minimum Ct value** | | | |
| 25-35 | 6.3±5.5 (1.0-21.0) | 0.5 (0.1-2.1) | |
| <25 | 6.7±3.1 (2.0-15.0) | Reference | |
| **Serostatus at end** | | | |
| Seronegative | 6.1±3.0 (2.0-14.0) | Reference | |
| Seropositive | 6.9±4.6 (1.0-21.0) | 0.6 (0.1-2.5) | |

| Characteristic | Serial interval, days, Mean ± SD (Range) | Univariate, HR* (95%CI) | Multivariable, aHR (95%CI) |
|---|---|---|---|
| Unknown | 5.0±2.0 (3.0-7.0) | 1.7 (0.1-22.8) | |
| **SARS-CoV-2 variant** | | | |
| non-Alpha/Beta/Delta | 8.2±2.3 (6.0-12.0) | 0.2 (0.0-1.0) | |
| Alpha | 3.6±1.2 (2.0-5.0) | 7.1 (0.7-73.2) | |
| Beta | 6.1±4.6 (1.0-21.0) | Reference | |
| Delta | 8.2±4.1 (2.0-15.0) | 0.4 (0.1-2.7) | |
| **Contact characteristics** | | | |
| **Age (years)** | | | |
| <5 | 5.5±0.7 (5.0-6.0) | 1.9 (0.1-26.0) | 0.5 (0.0-11.9) |
| 5-12 | 5.4±3.4 (1.0-10.0) | 0.9 (0.2-3.4) | 0.8 (0.2-2.9) |
| 13-17 | 6.3±3.5 (2.0-12.0) | 0.4 (0.1-1.3) | 0.3 (0.1-1.1) |
| 18-34 | 4.4±2.6 (2.0-9.0) | Reference | Reference |
| 35-59 | 7.5±4.7 (2.0-20.0) | **0.3 (0.1-1.1)** | **0.3 (0.1-0.9)** |
| ≥60 | 8.3±4.8 (3.0-21.0) | **0.3 (0.1-1.0)** | **0.2 (0.0-0.8)** |
| **Sex** | | | |
| Male | 6.2±4.6 (1.0-20.0) | Reference | |
| Female | 6.7±3.8 (2.0-21.0) | 0.8 (0.3-1.9) | |
| **HIV** | | | |
| HIV-uninfected | 5.4±4.0 (1.0-21.0) | Reference | Reference |
| HIV-infected | 11.1±5.6 (3.0-20.0) | **0.2 (0.0-1.0)** | **0.1 (0.0-0.8)** |
| Unknown | 6.6±2.8 (2.0-12.0) | 0.7 (0.2-2.3) | 1.0 (0.3-3.4) |
| **Underlying illness‡** | | | |
| No | 6.3±4.3 (1.0-21.0) | 2.6 (0.7-9.4) | |
| Yes | 7.6±3.3 (2.0-14.0) | Reference | |
| **Body-mass index** | | | |
| Underweight | 6.5±5.7 (3.0-15.0) | 0.3 (0.1-1.8) | |
| Normal weight | 6.7±4.4 (1.0-20.0) | 0.5 (0.1-1.6) | |
| Overweight | 5.9±3.4 (2.0-14.0) | Reference | |
| Obese | 6.7±4.2 (2.0-21.0) | 0.5 (0.1-1.6) | |
| **Current cigarette smoke (self-reported)** | | | |
| No | 6.3±3.7 (1.0-21.0) | Reference | |
| Yes | 7.6±5.9 (3.0-20.0) | 0.5 (0.1-1.9) | |
| **Serostatus at baseline** | | | |
| Seronegative | 6.6±3.4 (1.0-15.0) | 1.3 (0.5-3.2) | |
| Seropositive | 6.1±4.3 (2.0-21.0) | Reference | |
| Unknown | 8.5±7.9 (2.0-20.0) | 0.6 (0.1-4.4) | |

*Estimated using Weibull accelerated failure time regression adjusted for clustering by site and household. Hazard ratio <1 corresponds to prolonged serial interval compared to reference group. †Immune suppressed defined as CD4+ T cell count <200 cells/ml. ‡Underlying illness: self-reported history of diabetes, hypertension, asthma, lung disease, heart disease, stroke, spinal cord injury, epilepsy, cancer, liver disease, renal disease, pre-maturity.

**Supplementary Table 5.** Factors associated with at least one SARS-CoV-2 positive sample with a Ct value for any target ≤25 in secondary cases, Klerksdorp and Soweto, South Africa, 2020-2021, (n=232).

| Characteristic | Ct ≤25, n/N (%) | Univariate, OR (95%CI) | Multivariable, aOR (95%CI) |
|---|---|---|---|
| **Site** | | | |
| Klerksdorp | 44/99 (44.4) | Reference | |
| Soweto | 62/133 (46.6) | 1.2 (0.4-3.5) | |
| **Age (years)** | | | |
| <5 | 2/5 (40.0) | 0.0 (0.0-0.0) | |
| 5-12 | 16/40 (40.0) | Reference | |
| 13-17 | 22/46 (47.8) | 0.0 (0.0-0.0) | |
| 18-34 | 21/58 (36.2) | 0.7 (0.0-9.6) | |
| 35-59 | 30/60 (50.0) | 1.1 (0.3-3.7) | |
| ≥60 | 15/23 (65.2) | **0.8 (0.2-2.5)** | |
| **Sex** | | | |
| Male | 44/86 (51.2) | 2.3 (1.0-5.4) | |
| Female | 62/146 (42.5) | Reference | |
| **HIV** | | | |
| HIV-infected | 11/16 (68.8) | 4.2 (0.8-23.1) | |
| HIV-uninfected | 33/88 (37.5) | Reference | |
| Unknown HIV-status | 62/128 (48.4) | 1.9 (0.8-4.8) | |
| **Underlying illness‡** | | | |
| No | 85/198 (42.9) | Reference | Reference |
| Yes | 21/33 (63.6) | **2.2 (0.7-6.8)** | **3.7 (1.1-12.9)** |
| Unknown | 0/1 (0.0) | Excluded | Excluded |
| **Body-mass index** | | | |
| Underweight | 8/15 (53.3) | 2.7 (0.5-15.8) | |
| Normal weight | 51/102 (50.0) | 1.8 (0.7-4.7) | |
| Overweight | 22/47 (46.8) | 2.2 (0.7-7.1) | |
| Obese | 25/67 (37.3) | Reference | |
| Unknown | 0/1 (0.0) | Excluded | |
| **Current cigarette smoke (self-reported)** | | | |
| No | 90/205 (43.9) | Reference | |
| Yes | 16/26 (61.5) | 4.1 (1.0-17.1) | |
| Unknown | 0/1 (0.0) | Excluded | |
| **Symptoms** | | | |
| No | 63/163 (38.7) | Reference | Reference |
| Yes | 43/69 (62.3) | **4.0 (1.6-10.3)** | **3.2 (1.2-8.8)** |
| **Serostatus at follow-up start** | | | |
| Seronegative | 75/122 (61.5) | **5.3 (2.3-12.2)** | **9.4 (3.7-23.5)** |
| Seropositive | 25/94 (26.6) | Reference | Reference |
| Unknown | 6/16 (37.5) | 2.9 (0.5-18.6) | 3.7 (0.6-23.9) |
| **Serostatus at follow-up end** | | | |
| Seronegative | 5/22 (22.7) | Reference | |
| Seropositive | 85/164 (51.8) | **10.3 (1.8-59.7)** | **18.2 (3.4-95.9)** |
| Unknown | 16/46 (34.8) | 5.0 (0.7-35.3) | 9.4 (1.6-56.7) |
| **SARS-CoV-2 variant** | | | |

| | | | |
|---|---|---|---|
| non-Alpha/Beta/Delta | 10/18 (55.6) | 5.2 (0.7-37.4) | 2.2 (0.3-13.8) |
| Alpha | 7/16 (43.8) | 0.9 (0.1-7.8) | 1.2 (0.2-9.4) |
| Beta | 57/144 (39.6) | Reference | Reference |
| Delta | 32/52 (61.5) | **4.0 (1.0-16.9)** | **4.6 (1.2-17.9)** |
| Unknown | 0/2 (0.0) | Excluded | Excluded |

OR – Odds ratio, aOR – Adjusted odds ratio. † Underlying illness: self-reported history of diabetes, hypertension, asthma, lung disease, heart disease, stroke, spinal cord injury, epilepsy, cancer, liver disease, renal disease, pre-maturity. Bold face font denotes p<0.05.

**Supplementary Table 6.** COVID-19 related hospitalisations (n=11) and deaths (n=2), Klerksdorp and Soweto, South Africa, 2020-2021.

| **Index/ Contact** | **Age group** | **Comorbidities** | **Outcome** | **Length of admission (days)** | **Admission diagnosis** | **Infecting variant** |
|---|---|---|---|---|---|---|
| Index | 35-59 | Hypertension | Discharged alive | 6 | Shortness of breath | Beta |
| Contact | ≥60 | None | Discharged alive | 15 | Pneumonia | Beta |
| Contact | 13-17 | Congenital heart disease | Discharged alive | 2 | Shortness of breath | Alpha |
| Index | ≥60 | None | Discharged alive | 3 | Shortness of breath | Beta |
| Index | ≥60 | Hypertension, diabetes | Discharged alive | 13 | Shortness of breath | Beta |
| Index | ≥60 | None | Discharged alive | 4 | Shortness of breath | Alpha |
| Index | 35-59 | Hypertension | Died | 30 | Lower respiratory tract infection | Unknown |
| Index | 35-59 | None | Discharged alive | 3 | Shortness of breath | Delta |
| Contact | ≥60 | Asthma | Died | 8 | Pneumonia | Delta |
| Contact | ≥60 | None | Discharged alive | 11 | Not available | non-Alpha/Beta /Delta |
| Contact | ≥60 | None | Discharged alive | 12 | Shortness of breath | Beta |

**Supplementary Table 7.** Factors associated with SARS-CoV-2 household transmission in index cases and acquisition in household contacts in sensitivity analysis when including individuals seropositive at baseline with no rRT-PCR infection during follow-up as susceptible, Klerksdorp and Soweto, South Africa, 2020-2021, (n=444).

| Characteristic | HCIR*, n/N (%) | Univariate, OR (95%CI) | Multivariable, aOR (95%CI) |
|---|---|---|---|
| **Index characteristics** | | | |
| **Site** | | | |
| Klerksdorp | 95/208 (45.7) | Reference | |
| Soweto | 125/236 (53.0) | 1.4 (0.7-2.7) | 1.2 (0.6-2.4) |
| **Age (years)** | | | |
| 18-34 | 54/131 (41.2) | Reference | Reference |
| 35-59 | 137/257 (53.3) | 1.9 (0.8-4.2) | **3.5 (1.5-8.6)** |
| ≥60 | 29/56 (51.8) | 1.4 (0.5-4.6) | 2.5 (0.7-8.3) |
| **Sex** | | | |
| Male | 64/133 (48.1) | 0.9 (0.4-2.0) | |
| Female | 156/311 (50.2) | Reference | |
| **HIV** | | | |
| HIV-infected | 51/103 (49.5) | 1.0 (0.4-2.3) | |
| HIV-uninfected | 163/331 (49.2) | Reference | |
| Unknown HIV-status | 6/10 (60.0) | 0.9 (0.1-14.3) | |
| **CD4+ T cell count** | | | |
| HIV-infected, not immune suppressed† | 30/60 (50.0) | Reference | Reference |
| HIV-infected, immune suppressed† | 8/18 (44.4) | 1.4 (0.2-11.2) | 1.8 (0.2-12.5) |
| HIV-infected, no CD4+ T cell count | 13/25 (52.0) | 1.0 (0.2-5.6) | 0.9 (0.2-5.0) |
| HIV-uninfected | 163/331 (49.2) | 1.1 (0.4-3.0) | 1.6 (0.5-4.6) |
| Unknown HIV-status | 6/10 (60.0) | 1.0 (0.1-17.4) | 0.5 (0.0-8.0) |
| **Underlying illness‡** | | | |
| No | 218/432 (50.5) | 3.3 (0.3-41.6) | |
| Yes | 2/7 (28.6) | Reference | |
| Unknown | 0/5 (0.0) | Excluded | |
| **Body-mass index** | | | |
| Underweight | 3/10 (30.0) | 0.3 (0.0-2.7) | **0.1 (0.0-1.2)** |
| Normal weight | 68/125 (54.4) | Reference | Reference |
| Overweight | 44/86 (51.2) | 0.9 (0.3-2.4) | 0.4 (0.1-1.1) |
| Obese | 105/218 (48.2) | 0.7 (0.3-1.7) | **0.4 (0.2-1.0)** |
| Unknown | 0/5 (0.0) | Excluded | Excluded |
| **Crowding** | | | |
| No | 56/116 (48.3) | 1.0 (0.5-2.2) | |
| Yes | 164/328 (50.0) | Reference | |
| **Child <5 years in home** | | | |
| No | 190/381 (49.9) | 1.4 (0.5-3.9) | |
| Yes | 30/63 (47.6) | Reference | |
| **Current cigarette smoke (self-reported)** | | | |
| No | 203/391 (51.9) | 2.6 (0.9-7.6) | |
| Yes | 17/48 (35.4) | Reference | |
| Unknown | 0/5 (0.0) | Excluded | |

| Characteristic | HCIR*, n/N (%) | Univariate, OR (95%CI) | Multivariable, aOR (95%CI) |
|---|---|---|---|
| **Episode duration** | | | |
| 1-10 days | 64/147 (43.5) | Reference | |
| 11-21 days | 53/115 (46.1) | 1.3 (0.5-3.2) | |
| >21 days | 101/176 (57.4) | **2.3 (1.0-5.3)** | |
| Positive at last visit | 2/6 (33.3) | 0.6 (0.0-10.7) | |
| **Fever** | | | |
| No | 110/218 (50.5) | Reference | |
| Yes | 110/226 (48.7) | 1.2 (0.6-2.3) | |
| **Minimum Ct value** | | | |
| >35 | 9/45 (20.0) | Reference | Reference |
| 25-35 | 86/174 (49.4) | **6.3 (1.8-22.4)** | **5.1 (1.3-20.1)** |
| <25 | 123/222 (55.4) | **8.2 (2.4-28.4)** | **5.4 (1.4-20.3)** |
| Unknown | 2/3 (66.7) | 14.6 (0.3-748.6) | 11.7 (0.2-594.6) |
| **Serostatus at follow-up end** | | | |
| Seronegative | 4/7 (57.1) | 0.7 (0.0-12.3) | |
| Seropositive | 182/343 (53.1) | Reference | |
| Unknown | 34/94 (36.2) | 0.4 (0.1-0.9) | |
| **SARS-CoV-2 variant** | | | |
| non-Alpha/Beta/Delta | 18/32 (56.3) | 1.4 (0.4-4.6) | 1.9 (0.6-6.2) |
| Alpha | 16/25 (64.0) | 1.8 (0.4-7.6) | 2.3 (0.5-10.0) |
| Beta | 144/302 (47.7) | Reference | Reference |
| Delta | 40/58 (69.0) | **2.6 (0.9-7.5)** | 3.0 (1.0-9.4) |
| Unknown | 2/27 (7.4) | 0.0 (0.0-0.3) | 0.1 (0.0-0.5) |
| **Contact characteristics** | | | |
| **Age (years)** | | | |
| <5 | 5/19 (26.3) | Reference | Reference |
| 5-12 | 40/80 (50.0) | 3.3 (0.8-13.6) | 1.8 (0.4-7.9) |
| 13-17 | 43/67 (64.2) | **9.7 (2.2-42.9)** | **6.1 (1.3-29.0)** |
| 18-34 | 56/124 (45.2) | **3.7 (0.9-14.8)** | 2.8 (0.6-12.0) |
| 35-59 | 56/109 (51.4) | **3.8 (0.9-15.6)** | 3.0 (0.7-13.3) |
| ≥60 | 20/45 (44.4) | **3.4 (0.7-15.8)** | 2.1 (0.4-10.3) |
| **Sex** | | | |
| Male | 83/188 (44.2) | Reference | |
| Female | 137/256 (53.5) | **1.7 (1.1-2.8)** | |
| **HIV** | | | |
| HIV-uninfected | 82/170 (48.2) | 1.3 (0.5-3.4) | |
| HIV-infected | 16/35 (45.7) | Reference | |
| HIV status unknown | 122/239 (51.1) | 1.1 (0.5-2.9) | |
| **Underlying illness‡** | | | |
| No | 213/425 (50.1) | Reference | |
| Yes | 6/10 (60.0) | 1.2 (0.2-6.6) | |
| Unknown | 1/9 (11.1) | 0.1 (0.0-0.7) | |
| **Body-mass index** | | | |
| Underweight | 15/29 (51.7) | 1.0 (0.4-2.8) | |
| Normal weight | 96/199 (48.2) | Reference | |
| Overweight | 44/100 (44.0) | 0.9 (0.5-1.7) | |

| Characteristic | HCIR*, n/N (%) | Univariate, OR (95%CI) | Multivariable, aOR (95%CI) |
|---|---|---|---|
| Obese | 64/107 (59.8) | **1.9 (1.0-3.6)** | |
| Unknown | 1/9 (11.1) | 0.1 (0.0-0.9) | |
| **Current cigarette smoke (self-reported)** | | | |
| No | 197/377 (52.3) | **2.7 (1.3-5.8)** | **3.1 (1.4-6.9)** |
| Yes | 22/62 (35.5) | Reference | Reference |
| Unknown | 1/5 (20.0) | 0.3 (0.0-6.0) | Excluded |
| **Sleep in same room as index** | | | |
| No | 150/316 (47.5) | Reference | |
| Yes | 70/128 (54.7) | 1.4 (0.8-2.3) | |
| **Index is main caregiver** | | | |
| No | 183/375 (48.8) | 1.1 (0.5-2.2) | |
| Yes | 37/69 (53.6) | Reference | |

OR – Odds ratio, aOR – Adjusted odds ratio. *Household cumulative infection risk (HCIR) excludes household with mixed SARS-CoV-2 variant clusters. †Immune suppressed defined as CD4+ T cell count <200 cells/ml. ‡Underlying illness: self-reported history of diabetes, hypertension, asthma, lung disease, heart disease, stroke, spinal cord injury, epilepsy, cancer, liver disease, renal disease, pre-maturity. Bold face font denotes p<0.05.

**Supplementary Table 8.** Factors associated with SARS-CoV-2 household transmission in index cases and acquisition in household contacts sensitivity analysis including only households where 65% of household members completed 65% of follow-up during the first three weeks, Klerksdorp and Soweto, South Africa, 2020-2021, (n=342).

| Characteristic | HCIR*, n/N (%) | Univariate, OR (95%CI) | Multivariable, aOR (95%CI) |
|---|---|---|---|
| **Index characteristics** | | | |
| **Site** | | | |
| Klerksdorp | 92/174 (52.9) | Reference | |
| Soweto | 108/168 (64.3) | 1.7 (0.8-3.7) | 1.2 (0.6-2.7) |
| **Age (years)** | | | |
| 18-34 | 48/94 (51.1) | Reference | Reference |
| 35-59 | 124/198 (62.6) | 1.7 (0.7-4.3) | **4.2 (1.6-11.3)** |
| ≥60 | 28/50 (56.0) | 1.3 (0.3-4.9) | 2.2 (0.6-7.9) |
| **Sex** | | | |
| Male | 59/103 (57.3) | 0.9 (0.4-2.1) | |
| Female | 141/239 (59.0) | Reference | |
| **HIV** | | | |
| HIV-infected | 42/75 (56.0) | 0.7 (0.3-1.9) | |
| HIV-uninfected | 152/258 (58.9) | Reference | |
| Unknown HIV-status | 6/9 (66.7) | 0.7 (0.0-12.6) | |
| **CD4+ T cell count** | | | |
| HIV-infected, immune not suppressed† | 26/47 (55.3) | Reference | Reference |
| HIV-infected, immune suppressed† | 6/10 (60.0) | 2.0 (0.1-26.9) | 2.1 (0.2-18.3) |
| HIV-infected, no CD4+ T cell count | 10/18 (55.6) | 0.9 (0.1-6.5) | 1.1 (0.2-5.8) |
| HIV-uninfected | 152/258 (58.9) | 1.5 (0.5-4.9) | 2.5 (0.8-7.7) |
| Unknown HIV-status | 6/9 (66.7) | 1.0 (0.0-22.3) | 0.5 (0.0-7.0) |
| **Underlying illness¶** | | | |
| No | 154/261 (59.0) | 1.0 (0.4-2.7) | |
| Yes | 46/79 (58.2) | Reference | |
| Unknown | 0/2 (0.0) | Excluded | |
| **Body-mass index** | | | |
| Underweight | 2/7 (28.6) | 0.2 (0.0-3.1) | **0.0 (0.0-0.6)** |
| Normal weight | 61/100 (61.0) | Reference | Reference |
| Overweight | 40/58 (69.0) | 1.5 (0.5-5.1) | 0.6 (0.2-2.0) |
| Obese | 97/175 (55.4) | 0.9 (0.3-2.1) | **0.4 (0.1-0.9)** |
| Unknown | 0/2 (0.0) | Excluded | Excluded |
| **Crowding** | | | |
| No | 48/74 (64.9) | 1.6 (0.6-4.1) | |
| Yes | 152/268 (56.7) | Reference | |
| **Child <5 years in home** | | | |
| No | 173/288 (60.1) | 2.2 (0.7-6.8) | |
| Yes | 27/54 (50.0) | Reference | |
| **Current cigarette smoke (self-reported)** | | | |
| No | 186/305 (61.0) | 3.2 (0.9-11.0) | |
| Yes | 14/35 (40.0) | Reference | |
| Unknown | 0/2 (0.0) | Excluded | |
| **Episode duration** | | | |
| 1-10 days | 58/116 (50.0) | Reference | |

| Characteristic | HCIR*, n/N (%) | Univariate, OR (95%CI) | Multivariable, aOR (95%CI) |
|---|---|---|---|
| 11-21 days | 48/84 (57.1) | 1.8 (0.7-5.2) | |
| >21 days | 92/136 (67.7) | **2.9 (1.1-7.3)** | |
| Positive at last visit | 2/5 (40.0) | 0.5 (0.0-11.3) | |
| **Fever** | | | |
| No | 103/176 (58.5) | Reference | |
| Yes | 97/166 (58.4) | 1.1 (0.5-2.4) | |
| **Minimum Ct value** | | | |
| >35 | 7/36 (19.4) | Reference | Reference |
| 25-35 | 79/136 (58.1) | **9.5 (2.4-37.5)** | **7.9 (2.0-32.3)** |
| <25 | 112/166 (67.5) | **15.0 (3.8-58.6)** | **5.9 (1.5-23.3)** |
| Unknown | 2/4 (50.0) | 4.6 (0.2-113.5) | 8.0 (0.3-186.0) |
| **Serostatus at follow-up end** | | | |
| Seronegative | 4/7 (57.1) | 0.4 (0.0-7.0) | |
| Seropositive | 174/284 (61.3) | Reference | |
| Unknown | 22/51 (43.1) | 0.3 (0.1-1.0) | |
| **SARS-CoV-2 variant** | | | |
| non-Alpha/Beta/Delta | 16/25 (64.0) | 1.2 (0.3-4.5) | 1.9 (0.5-6.5) |
| Alpha | 13/17 (76.5) | 2.3 (0.4-12.6) | 2.4 (0.5-12.8) |
| Beta | 130/227 (57.3) | Reference | Reference |
| Delta | 39/47 (83.0) | **4.1 (1.2-13.9)** | **5.2 (1.4-19.9)** |
| Unknown | 2/26 (7.7) | 0.0 (0.0-0.2) | 0.0 (0.0-0.3) |
| **Contact characteristics** | | | |
| **Age (years)** | | | |
| <5 | 5/16 (31.3) | Reference | Reference |
| 5-12 | 37/71 (52.1) | 2.8 (0.6-13.2) | 1.5 (0.3-6.5) |
| 13-17 | 41/58 (70.7) | **11.7 (2.3-60.4)** | **6.5 (1.3-32.6)** |
| 18-34 | 47/79 (59.5) | **5.0 (1.1-23.3)** | **4.3 (0.9-20.0)** |
| 35-59 | 53/87 (60.9) | **4.3 (0.9-19.7)** | 3.7 (0.8-17.2) |
| ≥60 | 17/31 (54.8) | **4.2 (0.7-24.0)** | 2.5 (0.4-13.7) |
| **Sex** | | | |
| Male | 75/149 (50.3) | Reference | |
| Female | 125/193 (64.8) | **2.3 (1.3-4.2)** | |
| **HIV** | | | |
| HIV-uninfected | 80/146 (54.8) | 1.4 (0.4-4.4) | |
| HIV-infected | 14/26 (53.9) | Reference | |
| HIV status unknown | 106/170 (62.4) | 1.6 (0.5-4.8) | |
| **Underlying illness¶** | | | |
| No | 171/299 (57.2) | Reference | |
| Yes | 28/39 (71.8) | 3.2 (1.1-9.0) | |
| Unknown | 1/4 (25.0) | 0.2 (0.0-5.4) | |
| **Body-mass index** | | | |
| Underweight | 13/24 (54.2) | 0.9 (0.3-2.7) | |
| Normal weight | 88/166 (53.0) | Reference | |
| Overweight | 42/72 (58.3) | 1.4 (0.7-3.0) | |
| Obese | 56/76 (73.7) | **3.9 (1.7-9.0)** | |
| Unknown | 1/4 (25.0) | 0.2 (0.0-6.7) | |
| **Current cigarette smoke (self-reported)** | | | |

| Characteristic | HCIR*, n/N (%) | Univariate, OR (95%CI) | Multivariable, aOR (95%CI) |
|---|---|---|---|
| No | 179/292 (61.3) | **3.2 (1.4-7.8)** | **4.2 (1.7-10.4)** |
| Yes | 20/47 (42.6) | Reference | Reference |
| Unknown | 1/3 (33.3) | 0.6 (0.0-23.7) | Excluded |
| **Sleep in same room as index** | | | |
| No | 135/242 (55.8) | Reference | |
| Yes | 65/100 (65.0) | 1.5 (0.8-2.8) | |
| **Index is main caregiver** | | | |
| No | 163/280 (58.2) | 1.2 (0.5-2.7) | |
| Yes | 37/62 (59.7) | Reference | |

OR – Odds ratio, aOR – Adjusted odds ratio. *Household cumulative infection risk (HCIR) excludes household members seropositive at baseline with no SARS-CoV-2 detected on PCR during follow-up and households with mixed SARS-CoV-2 variant clusters. †Immune suppressed defined as CD4+ T cell count <200 cells/ml. ‡Underlying illness: self-reported history of diabetes, hypertension, asthma, lung disease, heart disease, stroke, spinal cord injury, epilepsy, cancer, liver disease, renal disease, pre-maturity. Bold face font denotes $p<0.05$.

**Supplementary Table 9.** Factors associated with SARS-CoV-2 episode duration in index cases sensitivity analysis including only households where 65% of household members completed 65% of follow-up during the first three weeks, Klerksdorp and Soweto, South Africa, 2020-2021, (n=117). HR – Hazard ratio*

| Characteristic | Episode duration, days, Mean ± SD (Range) | Univariate, HR* (95%CI) | Multivariable, aHR (95%CI) |
|---|---|---|---|
| **Site** | | | |
| Klerksdorp | 17.1±12.3 (4.0-45.0) | Reference | Reference |
| Soweto | 22.2±13.4 (3.0-47.0) | **0.7 (0.5-1.0)** | **0.5 (0.3-0.8)** |
| **Age (years)** | | | |
| 18-34 | 14.7±10.7 (4.0-41.0) | Reference | Reference |
| 35-59 | 19.8±13.6 (3.0-47.0) | 0.6 (0.4-0.9) | **0.3 (0.2-0.5)** |
| ≥60 | 27.9±10.7 (6.0-44.0) | **0.4 (0.2-0.7)** | **0.2 (0.1-0.4)** |
| **Sex** | | | |
| Male | 20.6±14.4 (3.0-47.0) | Reference | |
| Female | 19.2±12.5 (3.0-46.0) | 1.2 (0.8-1.8) | |
| **HIV** | | | |
| HIV-infected | 20.0±13.0 (3.0-45.0) | Reference | |
| HIV-uninfected | 19.6±13.2 (3.0-47.0) | 1.0 (0.6-1.5) | |
| Unknown HIV-status | 18.5±10.6 (11.0-26.0) | 1.3 (0.3-5.6) | |
| **CD4+ T cell count** | | | |
| HIV-infected, immune not suppressed† | 21.7±14.1 (3.0-45.0) | Reference | Reference |
| HIV-infected, immune suppressed† | 20.7±15.5 (8.0-38.0) | 1.2 (0.3-4.2) | 0.8 (0.2-3.6) |
| HIV-infected, no CD4+ T cell count | 15.3±9.3 (9.0-33.0) | 1.9 (0.7-5.0) | 0.8 (0.3-2.5) |
| HIV-uninfected | 19.6±13.2 (3.0-47.0) | 1.1 (0.6-2.0) | 0.8 (0.4-1.6) |
| Unknown HIV-status | 18.5±10.6 (11.0-26.0) | 1.5 (0.4-6.8) | 0.8 (0.2-4.2) |
| **Underlying illness¶** | | | |
| No | 19.4±13.0 (3.0-45.0) | Reference | |
| Yes | 20.7±13.4 (3.0-47.0) | 0.8 (0.5-1.3) | |
| Unknown | 9.0±0 (9.0-9.0) | 4.2 (1.0-17.8) | |
| **Body-mass index** | | | |
| Underweight | 13.5±6.4 (9.0-18.0) | **2.7 (0.6-11.5)** | 3.5 (0.7-18.2) |
| Normal weight | 21.6±14.0 (6.0-45.0) | Reference | Reference |
| Overweight | 20.5±15.6 (3.0-47.0) | 1.1 (0.6-1.8) | **2.0 (1.0-3.9)** |
| Obese | 18.6±11.6 (4.0-46.0) | 1.3 (0.8-2.0) | **2.2 (1.3-3.8)** |
| Unknown | 9.0±0 (9.0-9.0) | 4.5 (0.6-34.0) | 1.3 (0.2-11.3) |
| **Crowding** | | | |
| No | 17.6±11.9 (3.0-43.0) | Reference | |
| Yes | 20.5±13.5 (3.0-47.0) | 0.8 (0.5-1.2) | |
| **Child <5 years in home** | | | |
| No | 20.5±13.1 (3.0-47.0) | 0.6 (0.3-1.1) | |
| Yes | 13.9±11.3 (4.0-35.0) | Reference | |
| **Current cigarette smoke (self-reported)** | | | |
| No | 20.4±13.2 (3.0-47.0) | 0.6 (0.3-1.1) | |
| Yes | 14.7±11.3 (3.0-37.0) | Reference | |
| Unknown | 9.0±0 (9.0-9.0) | 2.7 (0.3-20.4) | |
| **Fever** | | | |
| No | 17.1±11.9 (3.0-46.0) | Reference | |

| Characteristic | Episode duration, days, Mean ± SD (Range) | Univariate, HR* (95%CI) | Multivariable, aHR (95%CI) |
|---|---|---|---|
| Yes | 22.3±13.8 (3.0-47.0) | **0.6 (0.4-0.9)** | |
| **Minimum Ct value** | | | |
| >35 | 11.3±7.0 (3.0-26.0) | Reference | Reference |
| 25-35 | 16.2±13.4 (3.0-47.0) | **0.5 (0.2-0.9)** | 0.5 (0.3-1.1) |
| <25 | 24.6±12.1 (7.0-44.0) | **0.3 (0.1-0.5)** | **0.3 (0.1-0.6)** |
| Unknown | 8.0±0 (8.0-8.0) | 2.2 (0.3-17.7) | 6.2 (0.7-53.4) |
| **Serostatus at follow-up end** | | | |
| Seronegative | 7.5±2.1 (6.0-9.0) | Reference | Reference |
| Seropositive | 21.1±13.3 (3.0-47.0) | **0.1 (0.0-0.5)** | **0.0 (0.0-0.2)** |
| Unknown | 13.2±9.7 (3.0-38.0) | 0.3 (0.1-1.2) | **0.1 (0.0-0.7)** |
| **SARS-CoV-2 variant** | | | |
| non-Alpha/Beta/Delta | 14.6±9.8 (7.0-38.0) | 2.1 (1.1-4.2) | **3.3 (1.5-6.9)** |
| Alpha | 23.8±14.9 (7.0-41.0) | 0.7 (0.3-1.8) | 0.6 (0.2-1.5) |
| Beta | 20.8±13.8 (3.0-47.0) | Reference | Reference |
| Delta | 21.8±11.8 (4.0-44.0) | 1.2 (0.6-2.2) | **2.2 (1.1-4.5)** |
| Unknown | 10.9±6.9 (5.0-28.0) | 2.8 (1.4-5.6) | 1.4 (0.6-3.2) |

*Estimated using Weibull accelerated failure time regression adjusted for clustering by site and household. Hazard ratio <1 corresponds to longer episode duration than reference group used. Excluded 8 index cases positive at last specimen collected. †Immune suppressed defined as CD4+ T cell count <200 cells/ml. ‡Underlying illness: self-reported history of diabetes, hypertension, asthma, lung disease, heart disease, stroke, spinal cord injury, epilepsy, cancer, liver disease, renal disease, pre-maturity. Bold face font denotes p<0.05.

**Supplementary Table 10.** Factors associated with serial interval of SARS-CoV-2 in cases with symptomatic illness sensitivity analysis including only households where 65% of household members completed 65% of follow-up during the first three weeks, Klerksdorp and Soweto, South Africa, 2020-2021, (n=60). HR – Hazard ratio*

| Characteristic | Serial interval, days, Mean ± SD (Range) | Univariate, HR* (95%CI) | Multivariable, aHR (95%CI) |
|---|---|---|---|
| **Index characteristics** | | | |
| **Site** | | | |
| Klerksdorp | 5.9±4.1 (1.0-21.0) | Reference | |
| Soweto | 7.5±4.0 (2.0-20.0) | 0.4 (0.1-1.5) | 0.3 (0.1-1.4) |
| **Age (years)** | | | |
| 18-34 | 6.6±4.6 (1.0-15.0) | Reference | |
| 35-59 | 6.6±3.6 (2.0-21.0) | 0.5 (0.1-2.7) | |
| ≥60 | 7.0±6.4 (2.0-20.0) | 0.9 (0.1-7.7) | |
| **Sex** | | | |
| Male | 6.6±4.7 (2.0-21.0) | 1.3 (0.3-5.6) | |
| Female | 6.7±3.9 (1.0-20.0) | Reference | |
| **HIV** | | | |
| HIV-infected | 11.7±7.6 (3.0-21.0) | 0.1 (0.0-0.6) | |
| HIV-uninfected | 6.1±3.2 (1.0-15.0) | Reference | |
| **Underlying illness‡** | | | |
| No | 7.1±4.4 (1.0-21.0) | Reference | |
| Yes | 17.0±5.5 (2.0-14.0) | 1.7 (0.4-6.8) | |
| **Body-mass index** | | | |
| Normal weight | 7.2±5.8 (2.0-21.0) | 1.3 (0.2-8.4) | |
| Overweight | 6.9±3.5 (2.0-15.0) | Reference | |
| Obese | 6.3±3.7 (1.0-20.0) | 1.3 (0.2-7.0) | |
| **Crowding** | | | |
| No | 5.8±4.1 (1.0-15.0) | Reference | |
| Yes | 7.1±4.1 (2.0-21.0) | 0.6 (0.1-2.3) | |
| **Child <5 years in home** | | | |
| No | 6.6±4.2 (1.0-21.0) | 0.4 (0.0-3.3) | |
| Yes | 6.9±3.6 (2.0-12.0) | Reference | |
| **Current cigarette smoke (self-reported)** | | | |
| No | 6.8±4.2 (1.0-21.0) | 0.2 (0.0-5.1) | |
| Yes | 4.3±0.5 (4.0-5.0) | Reference | |
| **Episode duration** | | | |
| 1-10 days | 7.2±3.9 (2.0-15.0) | 1.5 (0.2-9.1) | |
| 11-21 days | 7.4±4.6 (1.0-21.0) | Reference | |
| >21 days | 5.9±4.0 (2.0-20.0) | 3.7 (0.6-21.6) | |
| Unknown | | | |
| **Fever** | | | |
| No | 6.9±4.8 (1.0-20.0) | 1.5 (0.4-5.7) | |
| Yes | 6.5±3.7 (2.0-21.0) | Reference | |
| **Minimum Ct value** | | | |
| 25-35 | 6.6±5.7 (1.0-21.0) | 0.6 (0.2-2.5) | |
| <25 | 6.7±3.1 (2.0-15.0) | Reference | |
| **Serostatus at end** | | | |

| Characteristic | Serial interval, days, Mean ± SD (Range) | Univariate, HR* (95%CI) | Multivariable, aHR (95%CI) |
|---|---|---|---|
| Seronegative | 6.1±3.0 (2.0-14.0) | Reference | |
| Seropositive | 7.1±4.7 (1.0-21.0) | 0.5 (0.1-2.2) | |
| Unknown | 5.0±2.0 (3.0-7.0) | 1.7 (0.1-23.5) | |
| **SARS-CoV-2 variant** | | | |
| non-Alpha/Beta/Delta | 11.0±8.2 (6.0-12.0) | 0.2 (0.0-0.9) | |
| Alpha | 4.2±0.8 (3.0-5.0) | 2.5 (0.2-30.5) | |
| Beta | 6.0±4.7 (1.0-21.0) | Reference | |
| Delta | 8.2±4.1 (2.0-15.0) | 0.4 (0.0-2.5) | |
| **Contact characteristics** | | | |
| **Age (years)** | | | |
| <5 | 5.5±0.7 (5.0-6.0) | 1.9 (0.1-25.3) | 0.5 (0.0-12.1) |
| 5-12 | 6.0±3.3 (1.0-10.0) | 0.8 (0.2-3.6) | 0.7 (0.2-3.0) |
| 13-17 | 6.3±3.5 (2.0-12.0) | 0.4 (0.1-1.4) | 0.3 (0.1-1.1) |
| 18-34 | 4.6±2.6 (2.0-9.0) | Reference | Reference |
| 35-59 | 7.5±4.7 (2.0-20.0) | **0.4 (0.1-1.2)** | **0.3 (0.1-1.0)** |
| ≥60 | 8.2±5.0 (3.0-21.0) | 0.3 (0.1-1.2) | **0.2 (0.1-1.0)** |
| **Sex** | | | |
| Male | 6.2±4.6 (1.0-20.0) | Reference | |
| Female | 6.9±3.8 (2.0-21.0) | 0.8 (0.3-1.9) | |
| **HIV** | | | |
| HIV-uninfected | 5.5±4.0 (1.0-21.0) | Reference | Reference |
| HIV-infected | 11.1±5.6 (3.0-20.0) | **0.2 (0.0-0.9)** | **0.2 (0.0-0.8)** |
| Unknown | 6.7±2.7 (2.0-12.0) | 0.7 (0.2-2.4) | 1.1 (0.3-4.2) |
| **Underlying illness‡** | | | |
| No | 6.4±4.3 (1.0-21.0) | 2.1 (0.6-7.6) | |
| Yes | 7.5±3.5 (2.0-14.0) | Reference | |
| **Body-mass index** | | | |
| Underweight | 6.5±5.7 (3.0-15.0) | 0.3 (0.0-1.6) | |
| Normal weight | 6.7±4.4 (1.0-20.0) | 0.4 (0.1-1.4) | |
| Overweight | 5.7±3.4 (2.0-14.0) | Reference | |
| Obese | 7.1±4.1 (2.0-21.0) | 0.4 (0.1-1.3) | |
| **Current cigarette smoke (self-reported)** | | | |
| No | 6.4±3.7 (1.0-21.0) | Reference | |
| Yes | 7.6±5.9 (3.0-20.0) | 0.6 (0.2-2.0) | |
| **Serostatus at baseline** | | | |
| Seronegative | 6.5±3.4 (1.0-15.0) | 1.3 (0.5-3.3) | |
| Seropositive | 6.3±4.3 (2.0-21.0) | Reference | |
| Unknown | 10.7±8.1 (5.0-20.0) | 0.1 (0.0-1.8) | |

*Estimated using Weibull accelerated failure time regression adjusted for clustering by site and household. Hazard ratio <1 corresponds to prolonged serial interval. †Immune suppressed defined as CD4+ T cell count <200 cells/ml. ‡Underlying illness: self-reported history of diabetes, hypertension, asthma, lung disease, heart disease, stroke, spinal cord injury, epilepsy, cancer, liver disease, renal disease, pre-maturity. Bold face font denotes p<0.05.

### Supplementary figures

a)

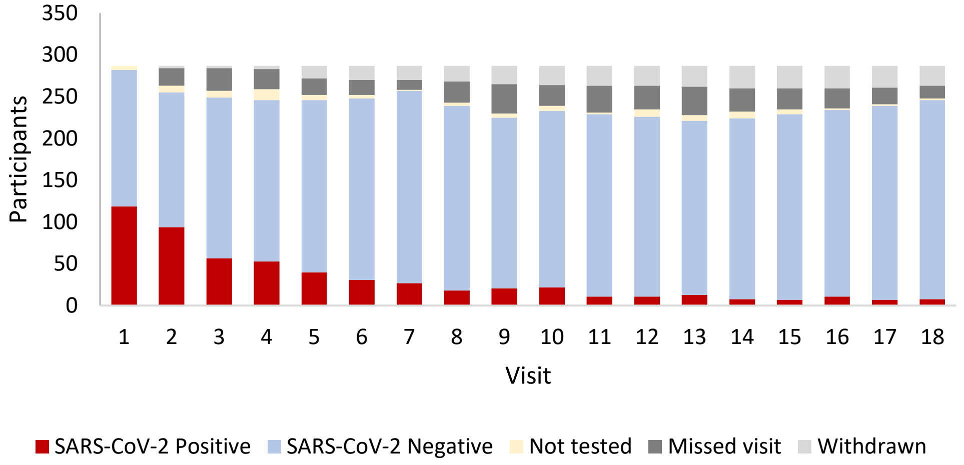

b)

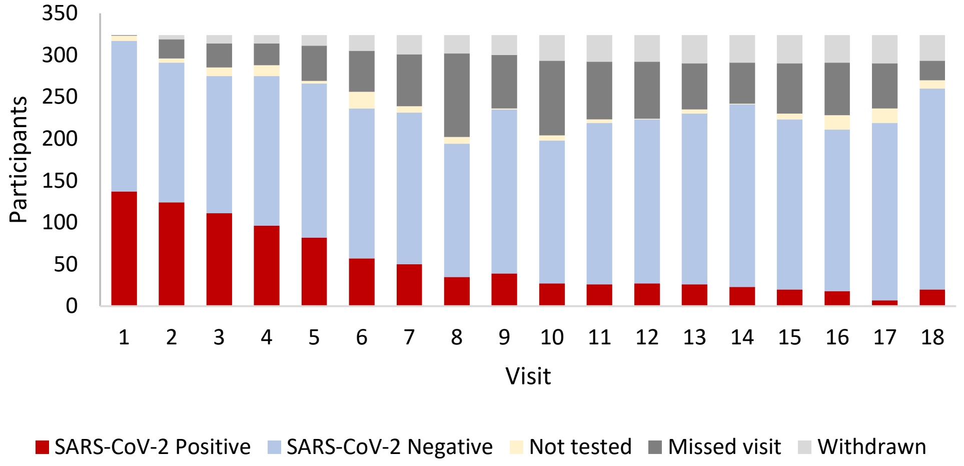

**Supplementary Figure 1.** SARS-CoV-2 positivity by follow-up visit for index cases and contacts*, a) Klerksdorp (n=287) and b) Soweto (n=325), South Africa, 2020-2021.

*Not tested: sample collected but not tested due to lost in transit, leakage or labelling error. Missed visit: visit not conducted as individual was not available.

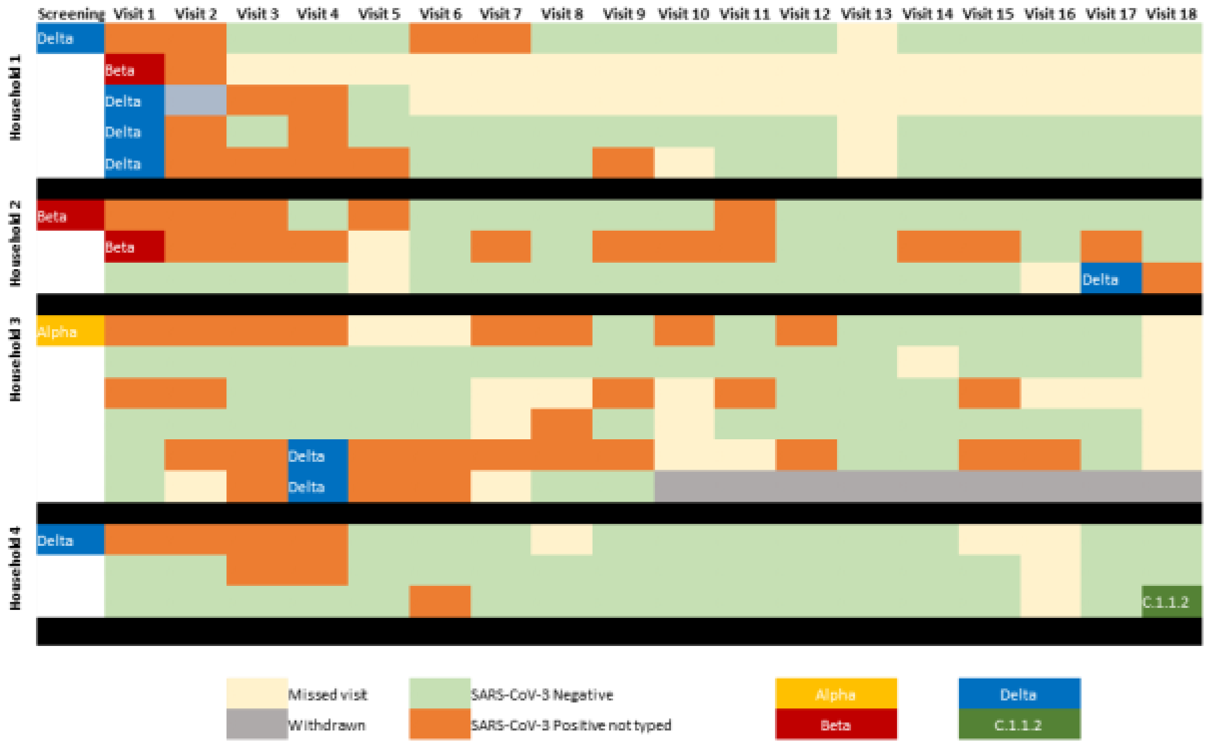

**Supplementary Figure 2.** Mixed SARS-CoV-2 clusters detected during follow-up, Klerksdorp and Soweto, South Africa, 2020-2021. Black line denotes household separation
